# Gender inequalities in the disruption of long-term life satisfaction trajectories during the COVID-19 pandemic and the role of time use: evidence from the 1970 British birth cohort

**DOI:** 10.1101/2023.11.15.23298585

**Authors:** Darío Moreno-Agostino, Jenny Chanfreau, Gemma Knowles, Alina Pelikh, Jayati Das-Munshi, George B. Ploubidis

**Affiliations:** ESRC Centre for Society and Mental Health, King’s College London, Melbourne House, 44-46 Aldwych, London WC2B 4LL, United Kingdom; Centre for Longitudinal Studies, UCL Social Research Institute, University College London; 55-59 Gordon Square, London WC1H 0NU, United Kingdom; Department of Sociology and Criminology, University of Sussex, Falmer, Brighton BN1 9RH, United Kingdom; Health Service and Population Research Department, Institute of Psychiatry, Psychology & Neuroscience, King’s College London, 16 De Crespigny Park, London, SE5 8AF; King’s College London, Department of Psychological Medicine, Institute of Psychiatry, Psychology & Neuroscience, 16 De Crespigny Park, London SE5 8AF, United Kingdom; South London and Maudsley NHS Trust, London, United Kingdom

## Abstract

**Background:** The COVID-19 pandemic has disproportionately impacted women’s mental health, although most evidence has focused on mental illbeing outcomes. Previous research suggests that gendered differences in time-use may explain this disparity, as women generally spend more time doing psychologically taxing activities than men. We investigated gender differences in the long-term trajectories of life satisfaction, how these were impacted during the pandemic, and the role of time-use differences in explaining gender inequalities.

**Methods:** We used data from 6766 (56.2% women) members of the 1970 British Cohort Study (BCS70), a nationally representative birth cohort of people born in Great Britain in 1970, who were alive and residing in the UK between May 2020 and March 2021. Life satisfaction was prospectively assessed between the ages of 26 (1996) and 51 (2021) using a single question with responses ranging from 0 (lowest) to 10 (highest). We analysed life satisfaction trajectories using piecewise latent growth curve models and investigated whether gender differences in the change in the life satisfaction trajectories with the pandemic were explained by self-reported time spent doing different paid and unpaid activities.

**Findings:** Women had consistently higher life satisfaction than men prior to the pandemic (Δ_intercept,unadjusted_=0.213 [95% CI: 0.087, 0.340], *p=*.001) and experienced a more accelerated decline with the pandemic onset (Δ_quad2,unadjusted_=−0.018 [−0.026, −0.011], *p*<.001). Time-use differences did not account for the more accelerated decrease in women’s life satisfaction levels with the pandemic (Δ_quad2,adjusted_=−0.016 [−0.031, −0.001], *p*=.035).

**Interpretation:** Our study shows pronounced gender inequalities in the impact of the pandemic on the long-term life satisfaction trajectories of adults in their 50s, with women losing their historical advantage over men. Self-reported time-use differences did not account for these inequalities, suggesting that they could be linked to other factors including the ‘mental load’ (invisible, unrecognised labour disproportionately undertook by women) or the menopausal transition.

## Introduction

There have been concerns that the COVID-19 pandemic has inequitably impacted the mental health of different groups within the population (Gibson et al., 2021; Sun et al., 2023). Certain groups, including women, have been disproportionately impacted, with evidence suggesting that pre-existing inequalities widened with the pandemic onset (Moreno-Agostino et al., 2023; Patel et al., 2022; Pierce et al., 2020). Multiple studies have tentatively proposed that gender inequalities in how people used their time may be partly responsible for this unequal impact, as women, on average, had a disproportionate burden of the additional caregiving responsibilities and household chores (Clark & Lepinteur, 2022; Hupkau & Petrongolo, 2020; Villadsen et al., 2020; Wettstein et al., 2022). However, empirical evidence for these hypotheses is lacking.

Although there is evidence on the impact of the pandemic on the long-term trajectories of mental illbeing outcomes such as psychological distress (Moreno-Agostino et al., 2023), such evidence is not available for mental wellbeing outcomes such as life satisfaction. These outcomes do not simply represent the other pole of the same axis as evidenced by studies showing that correlates of mental illbeing are not necessarily the same as correlates of mental wellbeing (Kinderman et al., 2015; Patalay & Fitzsimons, 2018). It is possible to experience high levels of distress *and* wellbeing, and the so-called “gender paradox” is precisely an example of this. This paradox refers to the finding that, despite being generally exposed to more disadvantage and experiencing higher levels of mental illbeing, women often have higher mental wellbeing levels than men in the population level (Blanchflower & Bryson, 2023; Helliwell et al., 2022; Joshanloo & Jovanovic, 2020; Zweig, 2015). However, recent evidence suggests that, during the pandemic, women may have lost that relative advantage in mental wellbeing (Blanchflower & Bryson, 2022; Office for National Statistics, 2022), without clear evidence on the potential mechanisms leading to that larger decline in women.

The aims of this study are twofold. First, to examine differences across women and men in the impact of the pandemic in their long-term trajectories of life satisfaction. This can provide relevant insights on whether there was a continuation of pre-existing trends or an unexpected disruption, as noted for psychological distress (Moreno-Agostino et al., 2023). Second, to formally investigate the potential role of time-use differences in these disparities. This can help reveal how gender norms and societal expectations shape individuals’ experiences and wellbeing during times of crisis and inform more gender-responsive policy responses to crises.

## Methods

### Sample and procedure

We used data from the 1970 British Cohort Study (BCS70), a birth cohort following the lives of around 17,000 people born in Great Britain in a single week in 1970 (Sullivan et al., 2022). Data on life satisfaction was collected at years 1996, 2000, 2004, 2012, and 2016 (corresponding to ages 26, 30, 34, 42, and 46). Cohort data was augmented using data from the COVID-19 survey (Brown et al., 2021), which collected data from the cohort members at three time points during the pandemic: May 2020, September/October 2020, and February/March 2021 (corresponding to ages 50, 50.5, and 51). Cohort members that took part in any of these COVID-19 survey waves were included in this study, leading to a total of 6,766 participants. Non-response weights were derived as part of the COVID-19 survey project in order to restore representativeness of BCS70 to its target population (adults born in 1970 in Britain, alive, and still residing in the UK during the pandemic), in line with the Centre for Longitudinal Studies’ missing data strategy (Mostafa et al., 2021). Details on the derivation of the non-response weights and their effectiveness to restore sample representativeness are available in the COVID-19 survey user guide (Brown et al., 2021). All participants provided informed consent.

### Measures

Life satisfaction was assessed by a single question with an 11-point response scale, with 0 and 10 representing the lowest and highest satisfaction, respectively. The specific wording of the questions varied slightly across the survey waves (see **Appendix S1**, Supplementary Material).

As part of the first and second COVID-19 survey waves, participants were asked to report the number of hours they usually spent doing different activities in a typical weekday since the outbreak (i.e., their time use). In this study, we focused on the time spent doing paid work (*working*), volunteering/doing unpaid work (*volunteering*), home-schooling children (*home-schooling*), doing other interactive activities with children (*caring:children*), caring for someone other than a child (*caring:other*), and doing housework (*housework*). Based on the distribution of responses to these variables (see **Appendix S2**, Supplementary Material), they were recoded into zero, up to eight, or more than eight hours (*working*); zero or more than zero hours (*volunteering*, *home-schooling*, *caring:children*, *caring:other*); or zero, up to one, or more than one hours (*housework*). Only information on time spent working was collected during the third COVID-19 survey wave, which was transformed into the same scale as the data from the first two COVID-19 survey waves.

In the three COVID-19 survey waves, participants were asked to report their financial situation compared to prior to the pandemic outbreak (“Much worse off”, “A little worse off”, “About the same”, “A little better off”, or “Much better off”), their work location (used to derive a variable capturing whether the person worked completely/partially from home or not), their keyworker status (yes/no). We derived a variable capturing whether the person was living with dependent children or young people aged ≤16 (Villadsen et al., 2020).

Complete data on sex assigned at birth was drawn from the birth sweep as information on gender was not consistently available for participants. Because of the mechanisms of action under study, which are likely reflective of different processes of socialisation and oppression, we believe that our observations reflect gender rather than sex inequalities, and therefore we refer to these as *gender* inequalities.

### Data analyses

#### Life course trajectories of life satisfaction

To address the first aim of the study, we used a latent growth curve modelling (LGCM) approach (Preacher, 2018), using observations from all data points from years 1996 to 2021. Models were unadjusted in order to avoid controlling for variables that may be on the pathway between gender and life satisfaction (Bartram, 2022). We analysed the life satisfaction trajectories across men and women separately, using a model comparison strategy to identify the best fitting functional form among a pre-specified set of options based on the inspection of the descriptive data. An intercepts-only/no change LGCM was estimated as a baseline model. We then estimated and compared the fit across a) polynomial trajectories (linear, quadratic, and cubic change), b) piecewise models with the knot at age 46 to accommodate the change in the trajectory shape with the pandemic onset, and c) a free/“latent basis” trajectory shape. All LGCM models were estimated using maximum likelihood with robust standard errors (MLR). Comparative fit of nested models was tested with the Satorra-Bentler Scaled robust chi-square difference test (Muthen, 2023), with a significant result indicating better fit of the most flexible model; whereas non-nested models were compared based on the information criteria, with lower information criteria values indicating better fit.

Once the optimal functional form was identified for each gender, a multiple group LGCM strategy was implemented in which increasing number of equality constraints across groups were specified to identify the most parsimonious version of the trajectories supported by the data. Then, equality of growth factors across genders was formally tested using Wald tests.

#### The role of time-use differences

To address the second aim of the study, we first introduced the time-use variables as predictors of changes in life satisfaction during the COVID-19 pandemic. We then further included financial situation, working from home, keyworker status, and living with any dependent child or young person aged ≤16 as potential confounders of the relationship between time use and life satisfaction. The rationale behind these analyses is that, if time use could account for the gender gap in life satisfaction during the pandemic, the gender differences in the growth parameters representing the change during the pandemic would be attenuated. As an additional exploratory analysis, gender differences in the impact of time spent in different activities on life satisfaction levels at each of the COVID-19 survey waves were tested using Wald tests.

#### Missing data

Inverse probability weighting was used to account for differential unit missingness and to restore the representativeness of the sample to the reference population, using the most recently available non-response weights for each individual (Brown et al., 2021). Item missingness was accounted for by using Full Information Maximum Likelihood (Enders & Bandalos, 2001). Both strategies were used simultaneously to render the missing-at-random (MAR) assumption more tenable.

#### Sensitivity analyses

Information on life satisfaction was collected across all waves using pen-and-paper (age 26 sweep) or computer (ages 30-51) self-reported questionnaires. A small portion (3.2%) of the participants in the last COVID-19 survey wave (February/March 2021) were interviewed by telephone to boost response. Since this subgroup or participants were allocated into this data collection mode not-at-random but based on their non-response to the online survey, this resulted in differences in some of the collected variables (including life satisfaction) potentially attributable to data collection mode (Brown et al., 2021). Since mode effects in life satisfaction measures have been long reported in the literature (Schwarz et al., 1991; Wavrock et al., 2023), we estimated the final unconditional and adjusted LGCMs including the interview mode as a covariate.

Since information on time use at the last time-point was limited, and to further ensure the temporal ordering of the variables in the analyses, we estimated an additional set of models in which the time-use variables and the confounders had a lagged (rather than concurrent) effect on the next life satisfaction assessment.

Data management was carried out in Stata version MP 17 (StataCorp, 2021). LGCM models were estimated in Mplus version 8 (Muthén & Muthén, 1998-2017). BCS70 deidentified data used for this study is available at the UK Data Service (UCL Centre for Longitudinal Studies, 2023). Example code of the unadjusted and adjusted multiple groups LGCMs is available on the Open Science Framework: https://osf.io/7esnw/.

## Results

Of the 6,766 participants who were alive and resided in the UK during the COVID-19 pandemic, 3,799 (56.2%) were women. A total of 42,804 observations were included, with a median of 7 contributed observations per individual (interquartile range = 5, 8). The mean and dispersion of life satisfaction levels over time between the ages of 26 and 51 (**Table 1**) was higher around midlife (between ages 34-42), seemingly dropping at an increasingly accelerated pace after that, coinciding with the pandemic onset. Mean life satisfaction levels were higher among women than men prior to the pandemic onset (at age 50), but this reversed during the pandemic. Visual depictions of the individual (unweighted) and mean (weighted) life satisfaction levels by gender are provided in **Appendix S3** and **Appendix S4**, respectively (Supplementary Material). A larger proportion of women reported spending time volunteering, caring for others, and doing more than one hour of housework; women were more likely to be keyworkers and less likely to work from home. A larger proportion of men reported working >8 hours, spending time taking care of children (beyond home-schooling), working from home, being in a better financial situation than before the outbreak, and living with dependent children or young people. During the first lockdown (age 50), a larger proportion of women reported spending time home-schooling children, but this was reversed before the reintroduction of lockdown measures in Autumn 2020 (age 50.5). Detailed information on the variables on time-use, financial situation, working-from-home, keyworker status, and living with dependent children or young people aged ≤16, collected during the COVID-19 pandemic, is provided in **Appendix S5** (Supplementary Material).

**Table 1.** Mean and dispersion of life satisfaction levels over time.

| Age / year | Men |  |  | Women |  |  | Overall |  |  |
| --- | --- | --- | --- | --- | --- | --- | --- | --- | --- |
|  | N<br>obs | M | SD | N<br>obs | M | SD | N<br>obs | M | SD |
| 26 / 1996 | 2117 | 7.21 | 1.81 | 2961 | 7.36 | 1.85 | 5078 | 7.30 | 1.83 |
| 30 / 2000 | 2522 | 7.35 | 1.65 | 3374 | 7.51 | 1.79 | 5896 | 7.44 | 1.73 |
| 34 / 2004 | 2376 | 7.49 | 1.59 | 3188 | 7.57 | 1.73 | 5564 | 7.54 | 1.67 |
| 42 / 2012 | 2628 | 7.46 | 1.80 | 3414 | 7.56 | 1.87 | 6042 | 7.52 | 1.84 |
| 46 / 2016 | 2586 | 7.37 | 1.75 | 3289 | 7.53 | 1.81 | 5875 | 7.46 | 1.79 |
| 50 / 2020 (May)* | 1599 | 7.25 | 1.90 | 2309 | 7.22 | 1.95 | 3908 | 7.23 | 1.93 |
| 50.5 / 2020 (Sep/Oct)* | 2102 | 7.17 | 1.93 | 2895 | 7.09 | 1.95 | 4997 | 7.13 | 1.94 |
| 51 / 2021 (Feb/Mar)* | 2312 | 6.93 | 1.96 | 3132 | 6.70 | 2.05 | 5444 | 6.80 | 2.02 |

*Note.* Unweighted results. M: mean; N obs: total number of observations; SD: standard deviation. Life satisfaction measure ranges from 0 (minimum) to 10 (maximum).
\* Data collected as part of the COVID-19 survey waves.

### Life course trajectories of life satisfaction

A piecewise model with two quadratic segments had the best fit in both women and men as evidenced by the model comparison strategy implemented to select the optimal functional form (fit indices of these models are provided in **Appendix S6**, Supplementary Material). The multiple group LCGM approach implemented to identify the most parsimonious model supported the inclusion of multiple equality constraints across genders in the long-term trajectories of life satisfaction (fit indices of these models are provided in **Appendix S7**, Supplementary Material). However, significant differences across groups were found in the intercepts –or starting points, located at age 23–, being 0.213 (95% CI 0.087, 0.340, *p* = 0.001) points higher among women; and in the quadratic change of the second segment – the accelerated decrease with the pandemic–, being −0.018 (95% CI −0.026, −0.011, *p* < 0.001) more accelerated among women. A visual representation of the resulting life satisfaction trajectories is provided in **Figure 1**. The results from the unconditional multiple group LGCM are included in **Table 2**.

**Figure 1.**
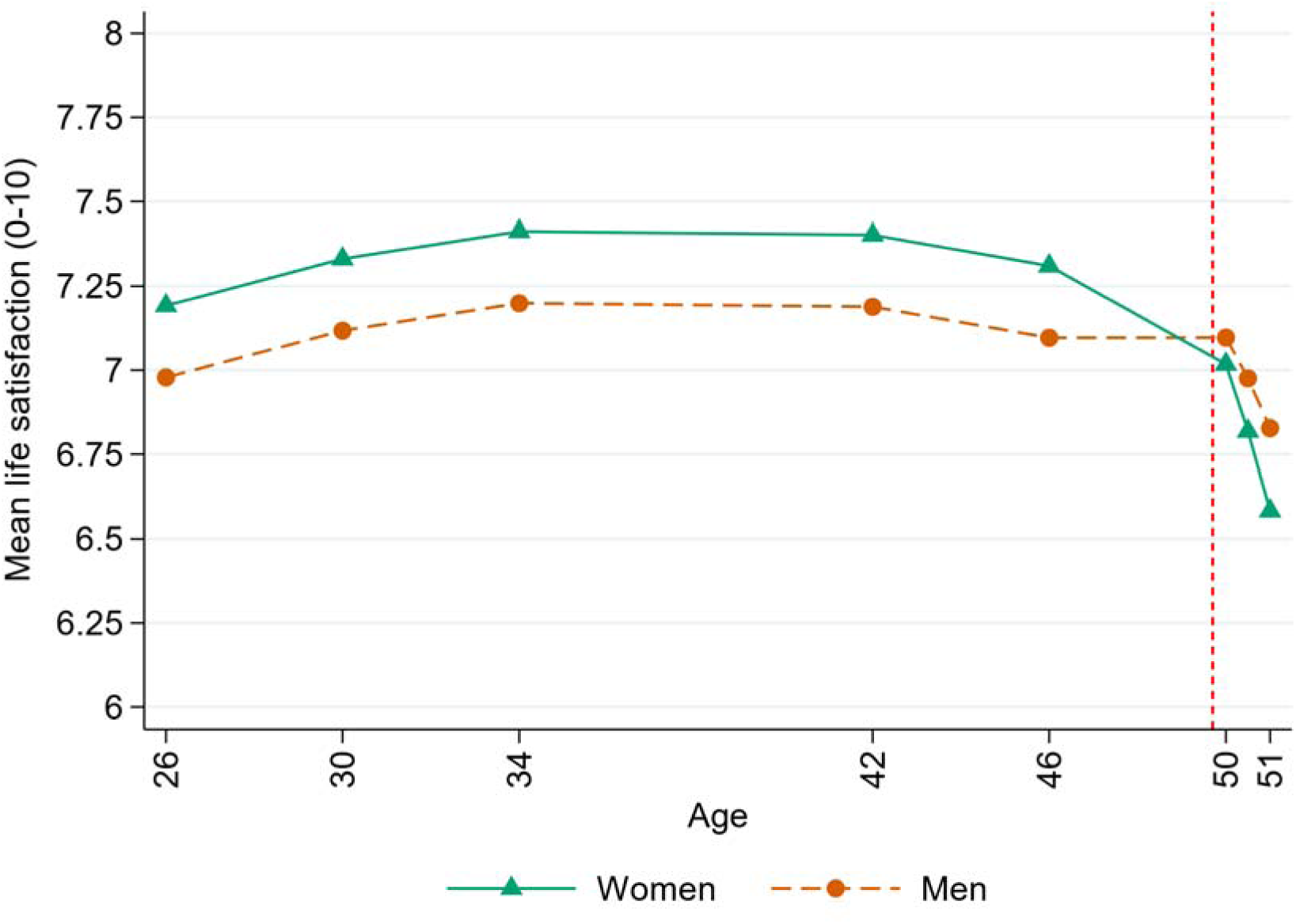
Unconditional life satisfaction trajectories across men and women (estimated, model-based means). The vertical red dashed line represents the COVID-19 pandemic onset.

**Table 2.** Results from the unconditional multiple group latent growth curve model (n=6,766).

|  | Women |  |  | Men |  |  |
| --- | --- | --- | --- | --- | --- | --- |
|  | Point estimate | 95% CI, LB | 95% CI, UB | Point estimate | 95% CI, LB | 95% CI, UB |
| **Means** |  |  |  |  |  |  |
| Intercept | 7.192 | 7.099 | 7.285 | 6.979 | 6.864 | 7.093 |
| Linear change, first segment | *0.042* | *0.026* | *0.058* | *0.042* | *0.026* | *0.058* |
| Quadratic change, first segment | -0.002 | -0.003 | -0.001 | -0.002 | -0.003 | -0.001 |
| Linear change, second segment | *0.216* | *0.119* | *0.312* | *0.216* | *0.119* | *0.312* |
| Quadratic change, second segment | -0.072 | -0.092 | -0.052 | -0.054 | -0.075 | -0.033 |
| **Variances** |  |  |  |  |  |  |
| Intercept | 1.697 | 1.464 | 1.930 | 1.697 | 1.464 | 1.930 |
| Linear change, first segment | *0.115* | *0.085* | *0.144* | *0.115* | *0.085* | *0.144* |
| Quadratic change, first segment | *0.000* |  |  | *0.000* |  |  |
| Linear change, second segment | 1.887 | 1.512 | 2.261 | 1.887 | 1.512 | 2.261 |
| Quadratic change, second segment | *0.000* |  |  | *0.000* |  |  |
| **Standardised covariances (correlations)** |  |  |  |  |  |  |
| Intercept x linear change, first segment | -0.347 | -0.472 | -0.221 | -0.321 | -0.451 | -0.191 |
| Intercept x linear change, second segment | -0.093 | -0.215 | 0.029 | -0.107 | -0.270 | 0.056 |
| Linear change first x second segment | -0.128 | -0.282 | 0.026 | -0.132 | -0.328 | 0.063 |
| **Standardised residual variances** |  |  |  |  |  |  |
| Age 26 | 0.557 | 0.500 | 0.613 | 0.551 | 0.489 | 0.613 |
| Age 30 | 0.564 | 0.518 | 0.611 | 0.516 | 0.456 | 0.576 |
| Age 34 | 0.548 | 0.502 | 0.593 | 0.481 | 0.419 | 0.543 |
| Age 42 | 0.544 | 0.483 | 0.605 | 0.508 | 0.452 | 0.563 |
| Age 46 | 0.399 | 0.325 | 0.473 | 0.383 | 0.300 | 0.465 |
| Age 50 | 0.414 | 0.342 | 0.487 | 0.271 | 0.221 | 0.322 |
| Age 50.5 | 0.299 | 0.257 | 0.341 | 0.283 | 0.222 | 0.345 |
| Age 51 | 0.338 | 0.287 | 0.390 | 0.275 | 0.222 | 0.329 |

Note. Models estimated using non-response weights and full information maximum likelihood under MLR estimation. Estimates in italics are constrained to be equal across groups. All estimates are significant with $p < 0.001$ , except the correlations involving the linear change in the second segment. CI: confidence interval; LB: lower bound; UB: upper bound.

The inclusion of the time-use variables resulted in a small attenuation of the differences in the accelerated change with the pandemic (represented by the quadratic term of the second segment of the piecewise latent growth model) and a large widening of the confidence interval [Δ_quad2_=−0.016 (−0.031, −0.001), *p* = 0.035]. Adding the covariates representing the financial situation, working from home and keyworker status, and the presence of dependent children or young people aged ≤16 in the household did not substantially change the differences across men and women in the accelerated decline in life satisfaction during the pandemic [Δ_quad2_=−0.022 (−0.038, −0.006), *p* = 0.008]. Results from the adjusted models are provided in **Appendix S8** (Supplementary Material).

Significant gender differences in the relationship between four time-use variables and life satisfaction were found in September/October 2020, a period without lockdown restrictions: working (doing paid work) was positively related to life satisfaction in women but negatively in men (Δ_working1-8h_=0.535 [0.156, 0.914], *p*=.006; Δ_working>8h_=0.560 [0.143, 0.978]; *p*=.009), whereas spending time caring for others (not children, Δ_caring1h_=−0.486 [−0.884, −0.087], *p*=.017) and doing housework (Δ_housework1h_=−0.555 [−0.956, −0.155], *p*=.007) was negatively related to life satisfaction in women but positively in men. Results from the Wald test conducted on the fully adjusted models are provided in **Appendix S9** (Supplementary Material).

Results were robust to the adjustment for potential mode effects (**Appendix S10**, Supplementary Material) and to the inclusion of lagged effects from the time-use variables and the confounders (**Appendix S11**, Supplementary Material).

## Discussion

Our study shows, for the first time, that the long-term trajectories of life satisfaction of British adults in their 50s significantly changed with the COVID-19 pandemic, reaching their lowest levels in up to 25 years of follow-up data. This decline was particularly salient among women, who lost their historical advantage over men reversing the pre-existing gender gap in life satisfaction, and our study suggests that self-reported differences in time use did not explain this differential impact. Although this historical advantage may seem small, its size (around 0.213-points higher life satisfaction in women than in men) is similar to the effect of not having health problems preventing engagement in activities that people of similar age (people aged 44-57) can normally do, as reported in a population-based study with data from multiple countries (Joshanloo & Jovanovic, 2021). Women not only lost that advantage, but their levels kept declining at a more rapid pace than men’s. Unlike what has been suggested in previous studies (Blanchflower & Bryson, 2022), our long-term longitudinal analysis does not suggest that this gendered change had started to occur prior to the pandemic (or at least not by 2016, the most recent pre-pandemic assessment). Our study aligns with recent research showing the disproportionate impact of the pandemic on women’s mental health outcomes in the UK (Milicev et al., 2022; Moreno-Agostino et al., 2023; Patel et al., 2022; Pierce et al., 2020; Sun et al., 2023) and complements existing literature on mental illbeing outcomes such as psychological distress (Moreno-Agostino et al., 2023) by placing that differential impact in the wider context of pre-existing long-term trajectories of life satisfaction. Unlike the evidence on mental illbeing, suggesting the widening of pre-existing inequalities (Moreno-Agostino et al., 2023), the present study focuses on a mental wellbeing outcome and suggests that the pandemic resulted in the emergence of a new, non-pre-existent inequality, with women born in 1970 losing their historical advantage over men in life satisfaction. Although this can be interpreted as a resolution for a “gender paradox” (Blanchflower & Bryson, 2022), with women now having both lower mental wellbeing and higher mental illbeing outcomes, it raises the important question of what potential mechanisms are driving this inequality (Patalay & Demkowicz, 2023).

In this regard, we engaged with that important question by investigating whether differences in how men and women used their time during the pandemic could account for at least part of this gap, but found inconclusive evidence. Our results are not consistent with previous studies suggesting that time-use differences might explain the gender gap in mental health (Milicev et al., 2022; Moreno-Agostino et al., 2023). Women’s higher engagement in activities typically associated with higher mental illbeing and lower mental wellbeing such as doing housework and caring for others (Moreno-Agostino et al., 2020) did not account for this disparity. A potential explanation for this is that is not the absolute time spent in the activity which may drive the gender gap in mental health, but the proportion of it that women do (Mencarini & Sironi, 2010). Additionally, society not only holds women accountable for these unpaid responsibilities, but women also tend to be responsible for coordinating and monitoring these activities even when not involved in their execution to a greater extent than men. This so-called ‘mental load’ has been proposed to have a ubiquitous toll on their cognitive and affective state as coordination and monitoring transcend time and place (Dean et al., 2022). Therefore, it is likely that this invisible, boundaryless labour plays a role in the observed life satisfaction inequalities during the pandemic. It is also possible that this ‘mental load’ went beyond the personal household and intensified during the pandemic, since adults in this are expected to care for their offspring and older parents in what has been called the ‘sandwich generation’ (Grundy & Henretta, 2006; Tosi & Grundy, 2018; Vlachantoni et al., 2020), potentially driving further gender inequalities. We also found that, during a period of reduced restrictions (September/October 2020), paid work, care work, and housework had different effects on life satisfaction for men and women. It may be that women, who were less likely to remain in the labour market (Dang & Viet Nguyen, 2021; Wielgoszewska et al., 2023), could derive more satisfaction from engaging in an activity (i.e., working) that had been more restricted in the initial stages of the pandemic. These results raise questions about whether there are systematic differences in the kind of activities that men and women report under general categories such as ‘doing housework’, which, in the administered questionnaire, comprised activities such as do-it-yourself (DIY) activities and household management chores, which may be related to higher and lower wellbeing, respectively (Hoang & Knabe, 2021). Overall, our study suggests that self-reported time-use differences did not explain the differential impact of the pandemic on women’s life satisfaction, even after accounting for potentially confounding factors such as the financial, occupational, and household composition situation. Future research on other potential mechanisms including the ability to socialise and perceive/receive social support (Joshanloo, 2018), the exposure to intimate partner violence and worries about personal safety (Kourti et al., 2023), the menopausal transition (Dennerstein et al., 2000), or the ‘mental load’ (Dean et al., 2022) is warranted to provide a deeper understanding of the mechanisms of the differential impact.

### Strengths and limitations

This study has several strengths and limitations. Unlike other studies conducted on convenience samples, we used data from a nationally representative cohort of people born in Britain in 1970, with data on the life satisfaction of the same individuals prospectively collected over 25 years, including during the COVID-19 pandemic. We accounted for differences in non-response by using appropriate methods to restore representativeness to the target population of adults alive and residing in the UK during the pandemic. By adopting a life course perspective, this study is the first to offer evidence on the impact of the pandemic on long-term life satisfaction trajectories in British adults, explicitly exploring potential mechanisms contributing to the differences found across women and men.

However, these findings must be interpreted with caution due to several limitations. Firstly, although the response options were nearly identical, slight variations in the wording of the life satisfaction questions over time may have introduced measurement error and influenced observed changes. Due to the single-item nature of the outcome, we could not empirically test its longitudinal measurement invariance. However, our findings align with other sources (Blanchflower & Bryson, 2022; Office for National Statistics, 2022), including cross-sectional evidence showing similar trends before and during the COVID-19 pandemic by gender in the UK, consistently using the same wording as that used in our study during the pandemic (Office for National Statistics, 2022). Secondly, there are several limitations regarding the measurement and operationalisation of time use. The reliance on questionnaire measures rather than diary ones may have increased recall bias and social desirability effects, and previous research in the UK has suggested that questionnaire-based estimates may underestimate the gender gap in housework participation (Kan, 2008). Additionally, as mentioned above, the broad categorisation of time-use activities hinders a more precise assessment of the specific activities encompassed in these reports. Furthermore, we could not adjust for differences in time-use at the final data collection point due to missing data in the corresponding survey wave. To mitigate this limitation, we included information on time spent working and the financial, occupational, and household composition covariates, and also explored the potential lagged effects of the time-use variables, but the gender gaps in the life satisfaction changes persisted. Our study covers data up to the first quarter of 2021, and the gender gap may have evolved subsequently, with repeated cross-sectional data suggesting that women’s and men’s life satisfaction levels may have converged by the second half of 2021 (Blanchflower & Bryson, 2022; Office for National Statistics, 2022). Future research using higher-resolution longitudinal data extending beyond the first year of the pandemic, along with less biased data collection methods such as ecological momentary assessments (Shiffman et al., 2008) or digital trackers, may yield additional insights into the role of time use in shaping the gender gap in mental health of the same individuals. While our study’s results may be generalisable to adults born in Britain in 1970 and residing in the UK, caution should be exercised when extending these findings to other segments of the UK population, such as migrants or different generations. For instance, younger generations may have been more likely to have younger children at home (Villadsen et al., 2020), with the impact of home-schooling and taking care of them being substantially different, potentially resulting in different gender inequalities. Finally, similar caution needs to be exercised when generalising these findings to other populations with different demographic, cultural, and political characteristics (Henrich et al., 2010).

## Conclusions

This study reveals a substantial decline in life satisfaction among adults in their fifties during the first year of the COVID-19 pandemic in Britain. These levels reached the lowest point observed across 25 years of follow-up data. Notably, the decline was most pronounced among women, who transitioned from having higher life satisfaction than men during most of their adult lives, to experiencing an even more rapid decline with the pandemic’s onset. Self-reported differences in time use did not account for the emergence of this new gender gap in life satisfaction. These results underscore the importance of ongoing monitoring of mental wellbeing in the population, especially as additional challenges (such as the cost-of-living crisis and underfunding of health, mental health, social care, and education services) continue to impact mental illbeing and wellbeing. It is crucial to prioritise the most disadvantaged individuals in these efforts. Further research is needed to understand the mechanisms driving the substantial decline in life satisfaction at the population level, particularly the more accelerated decline observed among women. These insights can inform the development of policies aimed at safeguarding and enhancing mental health during times of crisis.

## Data Availability

1970 British Cohort Study (BCS70) deidentified data used for this study is available at the UK Data Service.

https://beta.ukdataservice.ac.uk/datacatalogue/series/series?id=200001

## Acknowledgements

We would like to thank all individuals who participated in the BCS70 study for so generously giving up their time over so many years, and all the study team members for their tremendous efforts in collecting and managing the data.

## References

Bartram, D. (2022). The ‘Gender Life-Satisfaction/Depression Paradox’ Is an Artefact of Inappropriate Control Variables. Social Indicators Research, 164, 1061–1072.

Blanchflower, D.G., & Bryson, A. (2022). The Female Happiness Paradox (NBER Working Paper No. 29893). National Bureau of Economic Research: National Bureau of Economic Research.

Blanchflower, D.G., & Bryson, A. (2023). The Gender Well-being Gap (NBER Working Paper No. 31212). National Bureau of Economic Research: National Bureau of Economic Research.

Brown, M., Goodman, A., Peters, A., Ploubidis, G.B., Sanchez, A., Silverwood, R., et al. (2021). COVID-19 Survey in Five National Longitudinal Studies: Waves 1, 2 and 3 User Guide (Version 3). London: UCL Centre for Longitudinal Studies and MRC Unit for Lifelong Health and Ageing.

Clark, A.E., & Lepinteur, A. (2022). Pandemic Policy and Life Satisfaction in Europe. Rev Income Wealth, 68, 393–408.

Dang, H.H., & Viet Nguyen, C. (2021). Gender inequality during the COVID-19 pandemic: Income, expenditure, savings, and job loss. World Dev, 140, 105296.

Dean, L., Churchill, B., & Ruppanner, L. (2022). The mental load: building a deeper theoretical understanding of how cognitive and emotional labor overload women and mothers. Community Work & Family, 25, 13–29.

Dennerstein, L., Dudley, E., Guthrie, J., & Barrett-Connor, E. (2000). Life satisfaction, symptoms, and the menopausal transition. Medscape Women’s Health, 5, E4.

Enders, C.K., & Bandalos, D.L. (2001). The Relative Performance of Full Information Maximum Likelihood Estimation for Missing Data in Structural Equation Models. Structural equation modeling: a multidisciplinary journal, 8, 430–457.

Gibson, B., Schneider, J., Talamonti, D., & Forshaw, M. (2021). The Impact of Inequality on Mental Health Outcomes During the COVID-19 Pandemic: A Systematic Review. Canadian Psychology-Psychologie Canadienne, 62, 101–126.

Grundy, E., & Henretta, J.C. (2006). Between elderly parents and adult children: a new look at the intergenerational care provided by the ‘sandwich generation’. Ageing & Society, 26, 707–722.

Helliwell, J., Wang, S., Huang, H., & Norton, M. (2022). Happiness, Benevolence, and Trust During COVID-19 and Beyond. World Happiness Report 2022. New York: Sustainable Development Solutions Network.

Henrich, J., Heine, S.J., & Norenzayan, A. (2010). The weirdest people in the world? Behavioral and Brain Sciences, 33, 61–135.

Hoang, T.T.A., & Knabe, A. (2021). Time Use, Unemployment, and Well-Being: An Empirical Analysis Using British Time-Use Data. Journal of Happiness Studies, 22, 2525–2548.

Hupkau, C., & Petrongolo, B. (2020). Work, Care and Gender during the COVID-19 Crisis. *Fisc Stud*, 41, 623-651.

Joshanloo, M. (2018). Gender differences in the predictors of life satisfaction across 150 nations. Personality and Individual Differences, 135, 312–315.

Joshanloo, M., & Jovanovic, V. (2020). The relationship between gender and life satisfaction: analysis across demographic groups and global regions. Arch Womens Ment Health, 23, 331–338.

Joshanloo, M., & Jovanovic, V. (2021). Similarities and differences in predictors of life satisfaction across age groups: A 150-country study. Journal of Health Psychology, 26, 401–411.

Kan, M.Y. (2008). Measuring housework participation: The gap between “Stylised” questionnaire estimates and diary-based estimates. Social Indicators Research, 86, 381–400.

Kinderman, P., Tai, S., Pontin, E., Schwannauer, M., Jarman, I., & Lisboa, P. (2015). Causal and mediating factors for anxiety, depression and well-being. British Journal of Psychiatry, 206, 456–460.

Kourti, A., Stavridou, A., Panagouli, E., Psaltopoulou, T., Spiliopoulou, C., Tsolia, M., et al. (2023). Domestic Violence During the COVID-19 Pandemic: A Systematic Review. Trauma, Violence & Abuse, 24, 719–745.

Mencarini, L., & Sironi, M. (2010). Happiness, Housework and Gender Inequality in Europe. European Sociological Review, 28, 203–219.

Milicev, J., Qualter, P., Goodfellow, C., Inchley, J., Simpson, S.A., Leyland, A.H., et al. (2022). The prospective relationship between loneliness, life satisfaction and psychological distress before and during the COVID-19 pandemic in the UK. Z Gesundh Wiss, 1-15.

Moreno-Agostino, D., Fisher, H.L., Goodman, A., Hatch, S.L., Morgan, C., Richards, M., et al. (2023). Long-term psychological distress trajectories and the COVID-19 pandemic in three British birth cohorts: A multi-cohort study. PLoS Medicine, 20, e1004145.

Moreno-Agostino, D., Stone, A.A., Schneider, S., Koskinen, S., Leonardi, M., Naidoo, N., et al. (2020). Are retired people higher in experiential wellbeing than working older adults? A time use approach. Emotion, 20, 1411–1422.

Mostafa, T., Narayanan, M., Pongiglione, B., Dodgeon, B., Goodman, A., Silverwood, R.J., et al. (2021). Missing at random assumption made more plausible: evidence from the 1958 British birth cohort. Journal of Clinical Epidemiology, 136, 44–54.

Muthen, B. (2023). Chi-Square Difference Testing Using the Satorra-Bentler Scaled Chi-Square.

Muthén, L.K., & Muthén, B.O. (1998-2017). Mplus User’s Guide. Los Angeles, CA:. Office for National Statistics. (2022). Personal well-being in the UK: April 2011 to September 2021.

Patalay, P., & Demkowicz, O. (2023). Debate: Don’t mind the gap - why do we not care about the gender gap in common mental health difficulties? Child Adolesc Ment Health, 28, 341–343.

Patalay, P., & Fitzsimons, E. (2018). Development and predictors of mental ill-health and wellbeing from childhood to adolescence. Social Psychiatry and Psychiatric Epidemiology, 53, 1311–1323.

Patel, K., Robertson, E., Kwong, A.S.F., Griffith, G.J., Willan, K., Green, M.J., et al. (2022). Psychological distress before and during the COVID-19 pandemic among adults in the United Kingdom based on coordinated analyses of 11 longitudinal studies. JAMA Network Open, 5, e227629.

Pierce, M., Hope, H., Ford, T., Hatch, S., Hotopf, M., John, A., et al. (2020). Mental health before and during the COVID-19 pandemic: a longitudinal probability sample survey of the UK population. Lancet Psychiatry, 7, 883–892.

Preacher, K.J. (2018). Latent growth curve models. The reviewer’s guide to quantitative methods in the social sciences pp. 178-192): Routledge.

Schwarz, N., Strack, F., Hippler, H.-J., & Bishop, G. (1991). The impact of administration mode on response effects in survey measurement. Applied Cognitive Psychology, 5, 193–212.

Shiffman, S., Stone, A.A., & Hufford, M.R. (2008). Ecological momentary assessment. Annual Review of Clinical Psychology, 4, 1–32.

StataCorp. (2021). Stata Statistical Software: Release 17. College Station, TX: StataCorp LLC.

Sullivan, A., Brown, M., Hamer, M., & Ploubidis, G.B. (2022). Cohort Profile Update: The 1970 British Cohort Study (BCS70). International Journal of Epidemiology.

Sun, Y., Wu, Y., Fan, S., Dal Santo, T., Li, L., Jiang, X., et al. (2023). Comparison of mental health symptoms before and during the covid-19 pandemic: evidence from a systematic review and meta-analysis of 134 cohorts. BMJ, 380, e074224.

Tosi, M., & Grundy, E. (2018). Returns home by children and changes in parents’ well-being in Europe. Social Science and Medicine, 200, 99–106.

UCL Centre for Longitudinal Studies. (2023). 1970 British Cohort Study. [data series]. 9th Release. UK Data Service. SN: 200001.

Villadsen, A., Conti, G., & Fitzsimons, E. (2020). Parental involvement in home schooling and developmental play during lockdown-Initial findings from the COVID19 Survey in Five National Longitudinal Studies. London: UCL Centre for Longitudinal Studies.

Vlachantoni, A., Evandrou, M., Falkingham, J., & Gomez-Leon, M. (2020). Caught in the middle in mid-life: provision of care across multiple generations. Ageing & Society, 40, 1490–1510.

Wavrock, D., Schellenberg, G., & Boulet, C. (2023). Survey framing and mode effects in life satisfaction responses on Canadian social surveys. In S. Canada (Ed.), Economic and Social Reports: Statistics Canada.

Wettstein, M., Wahl, H.W., & Schlomann, A. (2022). The Impact of the COVID-19 Pandemic on Trajectories of Well-Being of Middle-Aged and older Adults: A Multidimensional and Multidirectional Perspective. J Happiness Stud, 1-28.

Wielgoszewska, B., Bryson, A., Costa Dias, M., Foliano, F., Joshi, H., & Wilkinson, D. (2023). Exploring the reasons for labour market gender inequality a year into the COVID-19 pandemic: evidence from the UK cohort studies. Longitudinal and life course studies: international journal, 14, 180–202.

Zweig, J.S. (2015). Are Women Happier than Men? Evidence from the Gallup World Poll. Journal of Happiness Studies, 16, 515–541.

