## Supplementary Material for "Gender inequalities in the disruption of long-term life satisfaction trajectories during the COVID-19 pandemic and the role of time use: evidence from the 1970 British birth cohort"

**Appendix S1. Life satisfaction question wording and mode of administration.**

| Age / year | Life satisfaction question wording | Mode of administration |
|---|---|---|
| 26 / 1996 | Here is a scale from 0 to 10. On it, “0” means that you are completely dissatisfied and “10” means that you are completely satisfied. Please tick the box with the number above it which shows how dissatisfied or satisfied you are about the way your life has turned out so far. | Pen-and-paper self-report questionnaire |
| 30 / 2000 | Here is a scale from 0-10 where ‘0’ means that you are completely dissatisfied and ‘10’ means that you are completely satisfied. Please enter the number which corresponds with how satisfied or dissatisfied you are about the way your life has turned out so far. | CASI |
| 34 / 2004 | Here is a scale from 0-10 where ‘0’ means that you are completely dissatisfied and ‘10’ means that you are completely satisfied. Please enter the number which corresponds with how satisfied or dissatisfied you are with the way life has turned out so far. | CASI |
| 42 / 2012 | Here is a scale from 0-10 where ‘0’ means that you are completely dissatisfied and ‘10’ means that you are completely satisfied. Please select the number which corresponds with how satisfied or dissatisfied you are with the way life has turned out so far. | CASI |
| 46 / 2016 | Here is a scale from 0-10 where ‘0’ means that you are completely dissatisfied and ‘10’ means that you are completely satisfied. Please select the number which corresponds with how satisfied or dissatisfied you are with the way life has turned out so far. | CASI |
| 50 / 2020 (May) | Overall, how satisfied are you with your life nowadays, where 0 means ‘not at all’ and 10 means ‘completely’? | CAWI |
| 50.5 / 2020 (September/October) | Overall, how satisfied are you with your life nowadays, where 0 means ‘not at all’ and 10 means ‘completely’? | CAWI |
| 51 / 2021 (February/March) | Overall, how satisfied are you with your life nowadays, where 0 means ‘not at all’ and 10 means ‘completely’? | CAWI + CATI |

*Note.* CASI: computer-assisted self-interview; CATI: computer-assisted telephone interview; CAWI: computer-assisted web-interview.

**Appendix S2. Distribution of responses to financial situation and time use variables.**

The figures below show the distribution of responses to financial situation and time use variables across the COVID-19 survey waves. Due to its reasonably symmetric distribution, financial situation was introduced in the models as a continuous variable in the models. Information on time use was only collected in waves 1 and 2 of the COVID-19 survey, although information on time spent working at the COVID-19 survey wave 3 was approximated using the self-report of hours spent working per week divided by five. A vertical red dashed line represents the values used as thresholds to recode the time use variables into categorical variables.

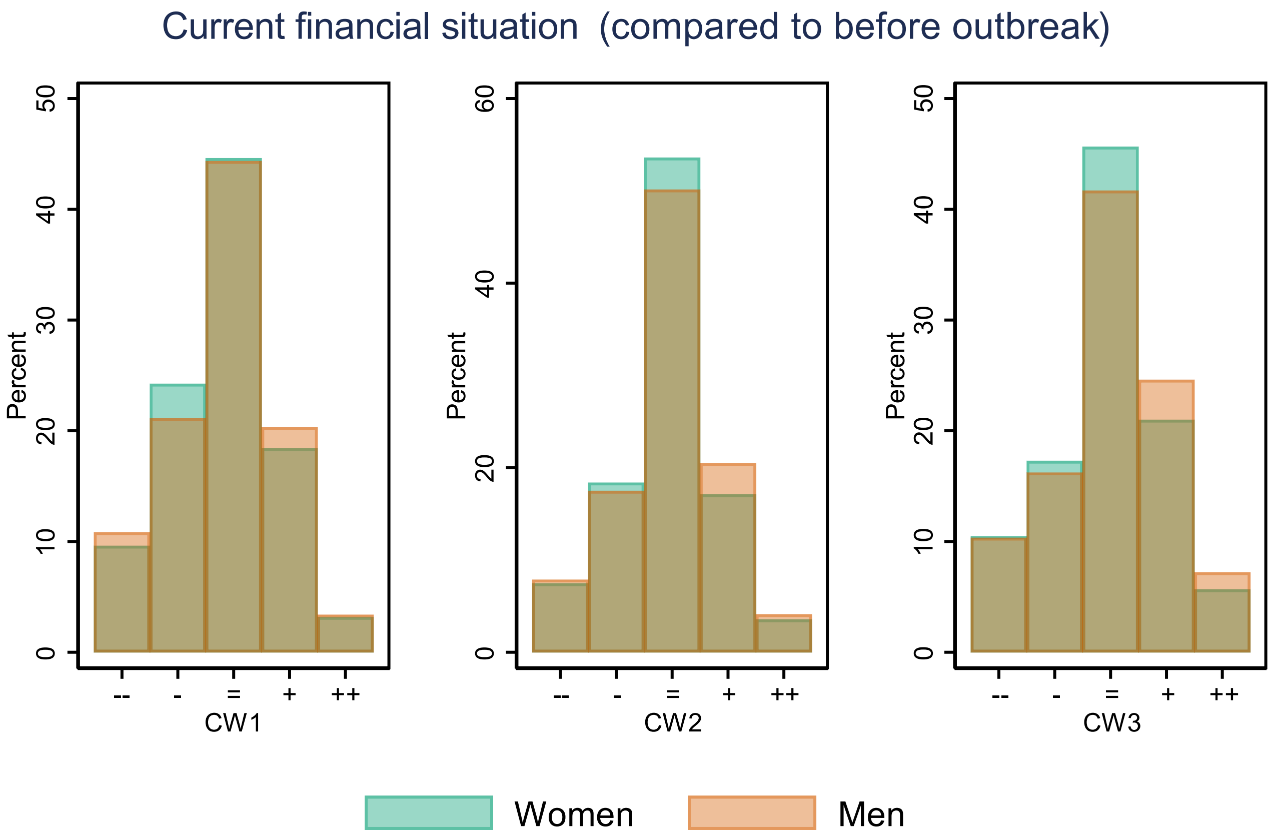

-- Much worse off; - A little worse off; = About the same; + A little better off; ++ Much better off.
CW1: COVID-19 survey wave 1 (May 2020); CW2: COVID-19 survey wave 2 (Sept-Oct 2020);
CW3: COVID-19 survey wave 3 (Feb-Mar 2021).

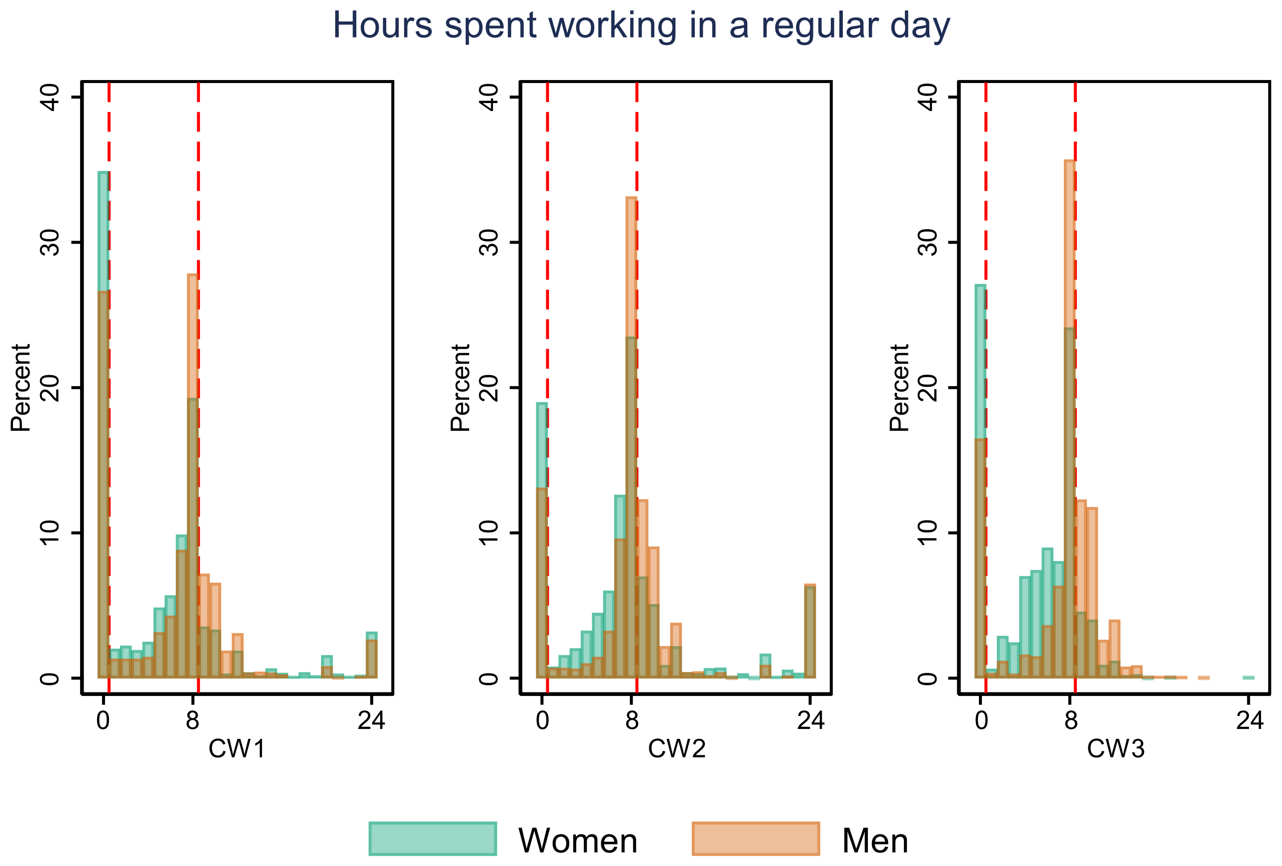

CW1: COVID-19 survey wave 1 (May 2020); CW2: COVID-19 survey wave 2 (Sept-Oct 2020);
CW3: COVID-19 survey wave 3 (Feb-Mar 2021).

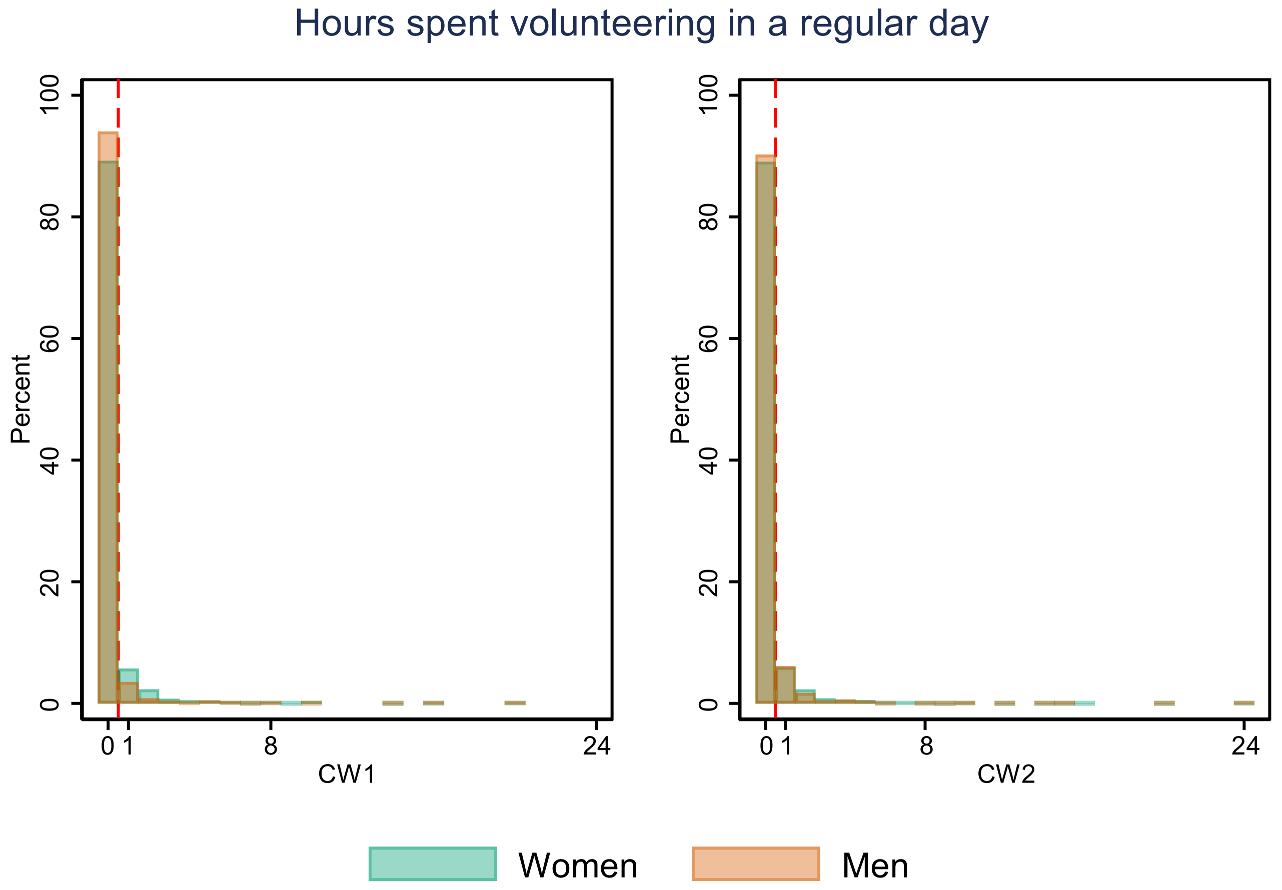

CW1: COVID-19 survey wave 1 (May 2020); CW2: COVID-19 survey wave 2 (Sept-Oct 2020)

Hours spent home-schooling children in a regular day

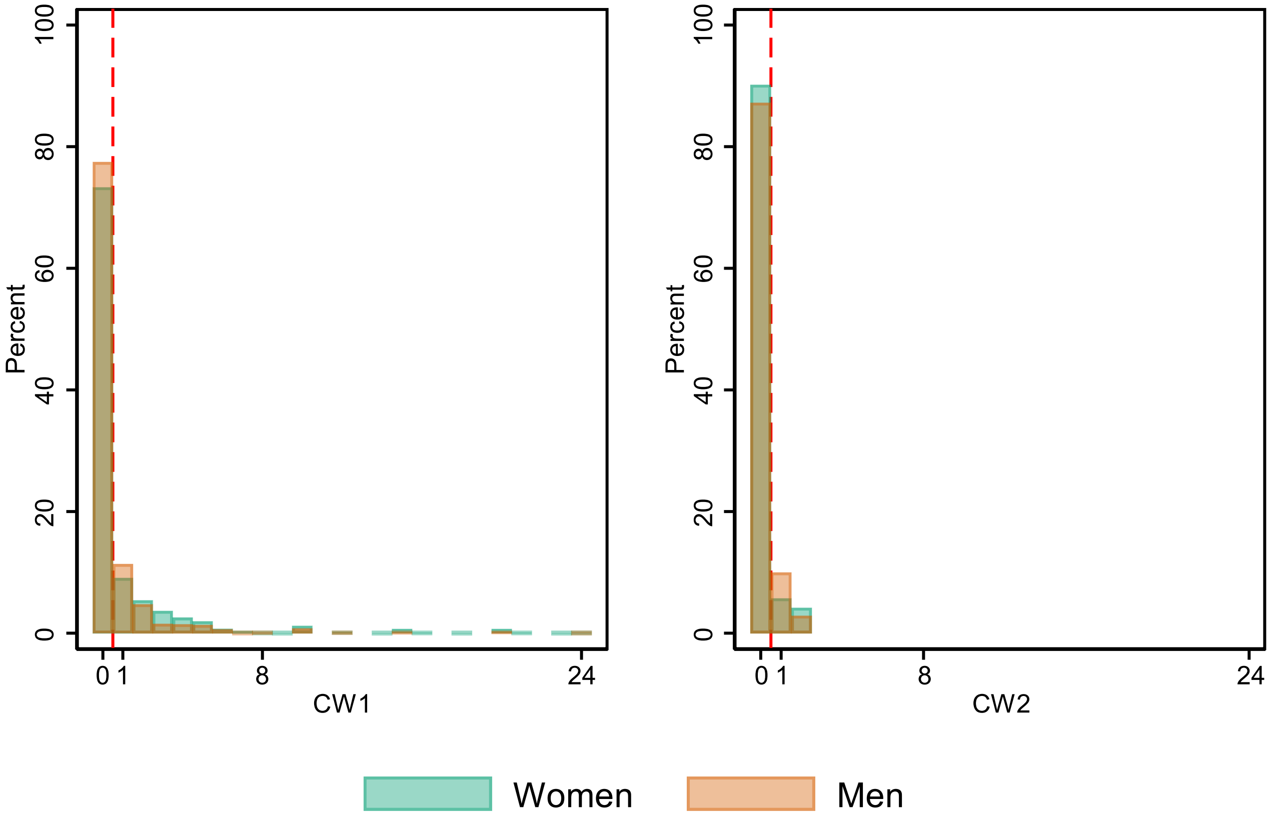

CW1: COVID-19 survey wave 1 (May 2020); CW2: COVID-19 survey wave 2 (Sept-Oct 2020)

Hours spent taking care of children (not homeschooling) in a regular day

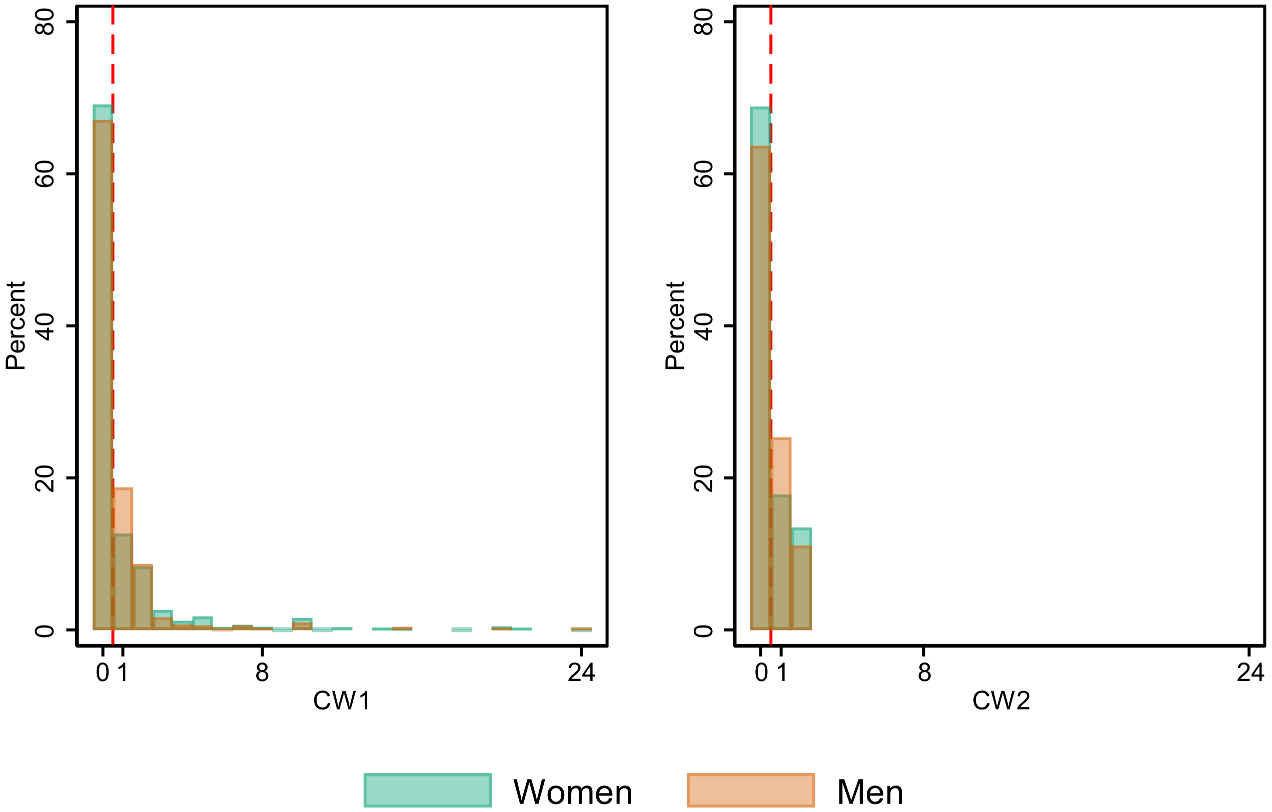

CW1: COVID-19 survey wave 1 (May 2020); CW2: COVID-19 survey wave 2 (Sept-Oct 2020)

Hours spent caring for others (not children) in a regular day

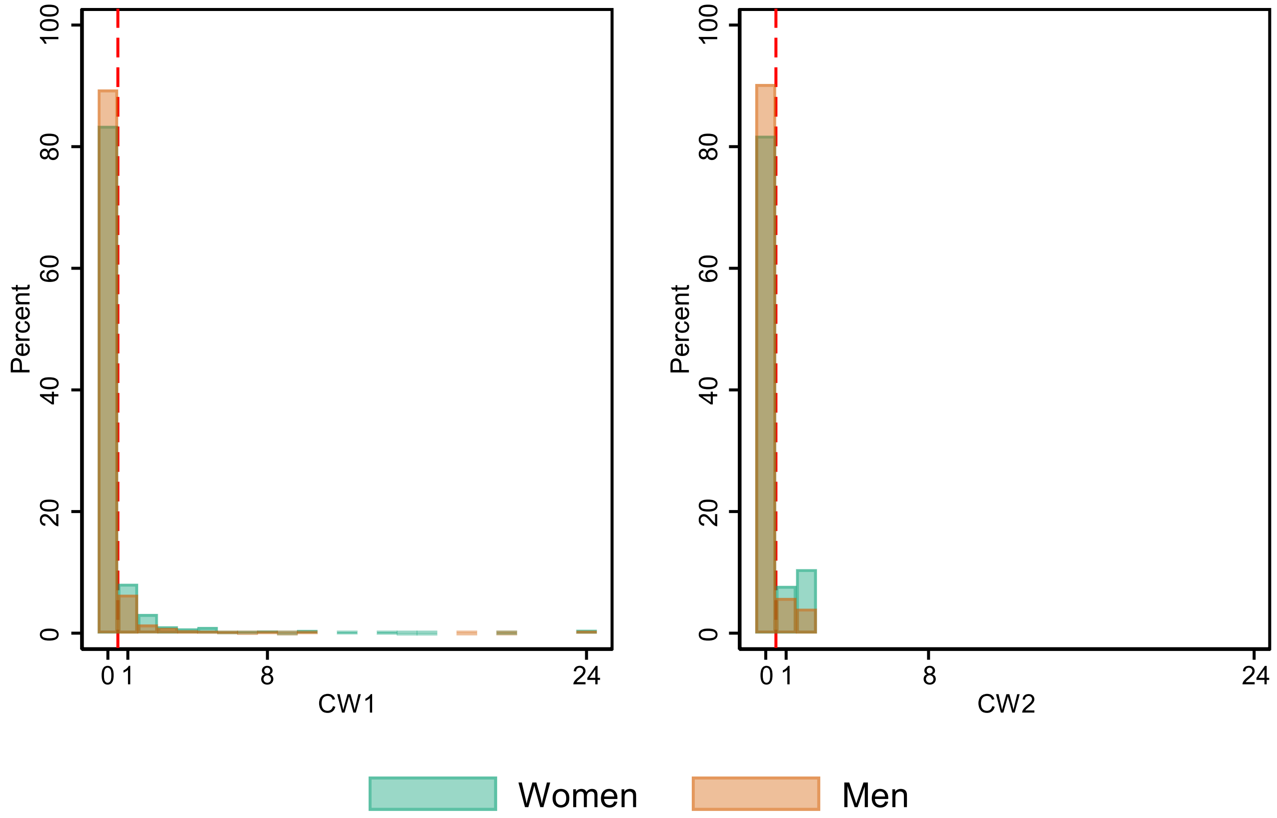

CW1: COVID-19 survey wave 1 (May 2020); CW2: COVID-19 survey wave 2 (Sept-Oct 2020)

Hours spent doing housework in a regular day

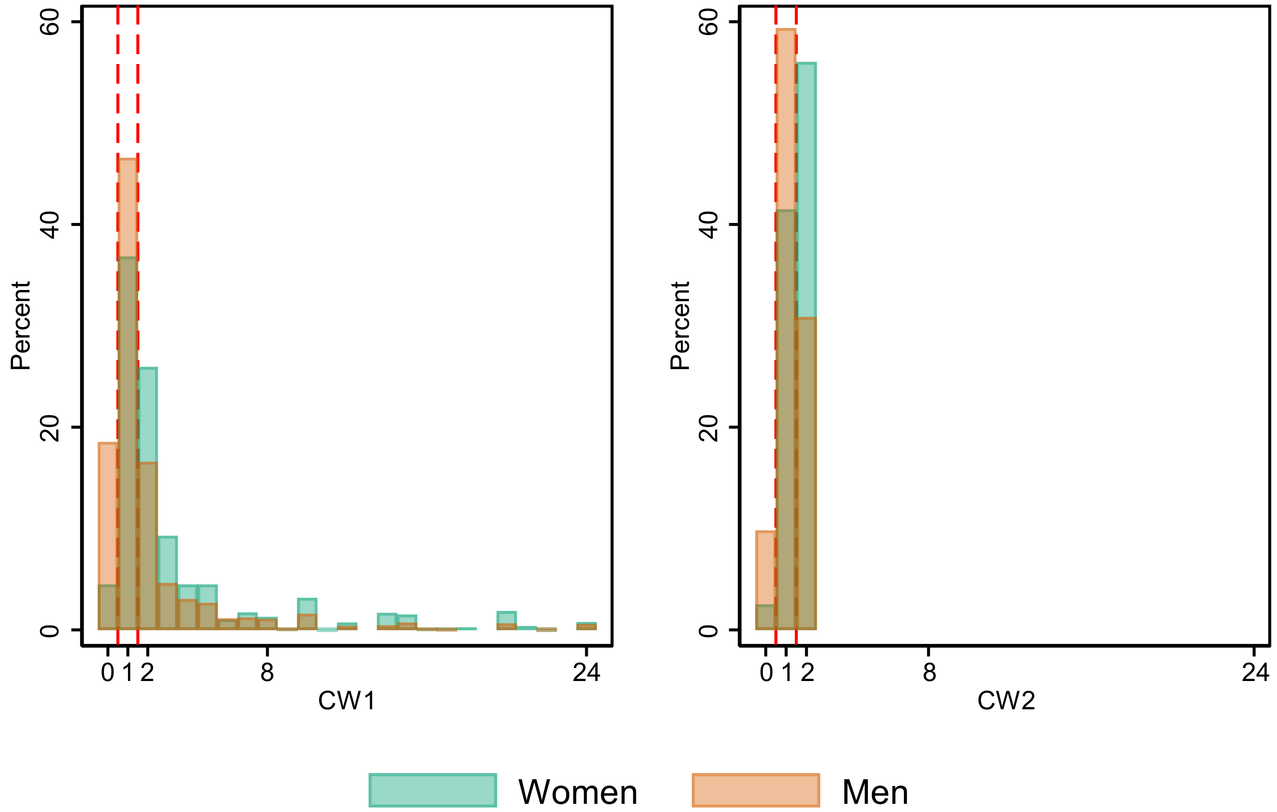

CW1: COVID-19 survey wave 1 (May 2020); CW2: COVID-19 survey wave 2 (Sept-Oct 2020)

**Appendix S3. Observed (unweighted) life satisfaction levels across a random (n=200) subset of cohort members over time.**

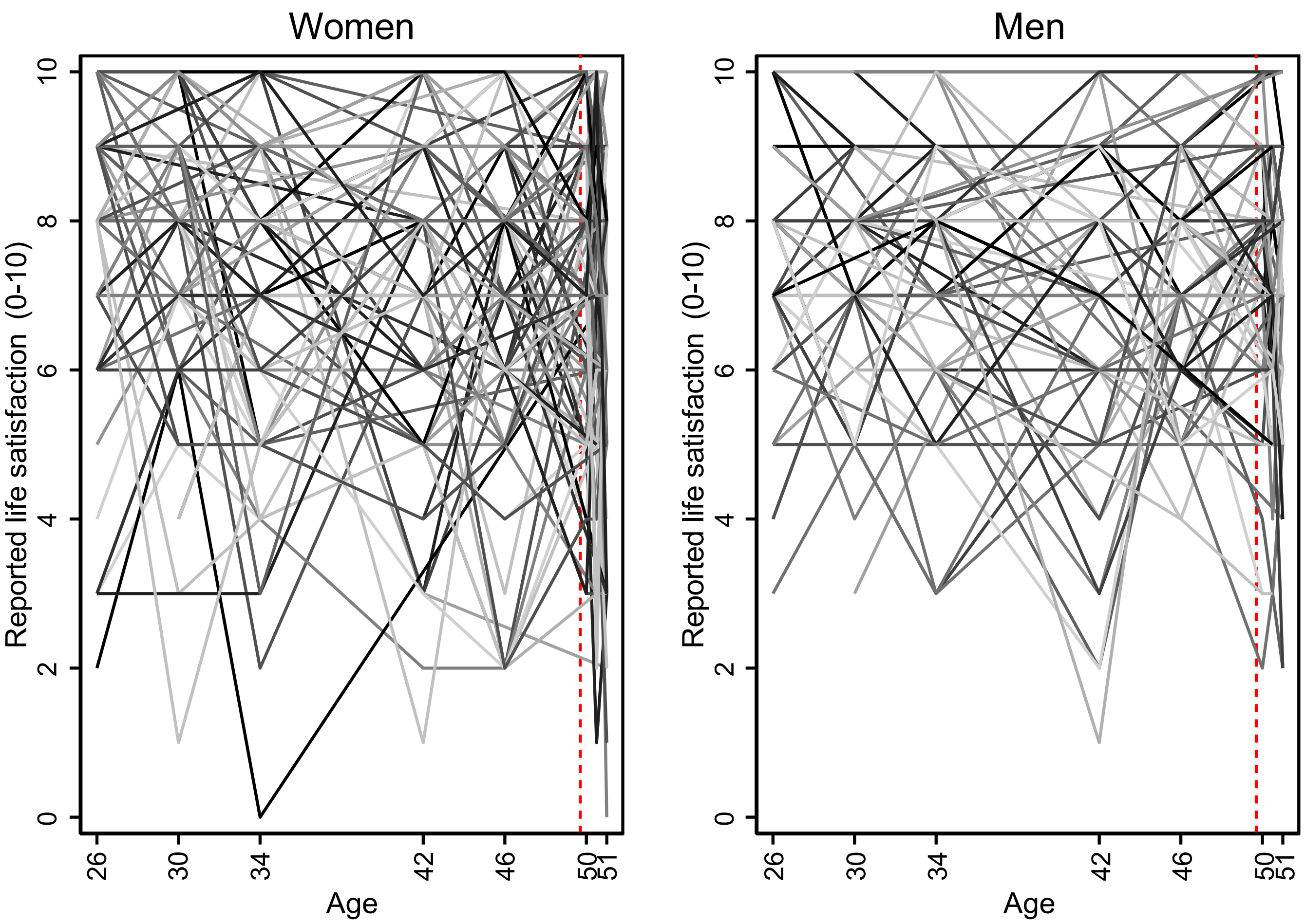

*Note.* The vertical red dashed line represents the pandemic onset.

**Appendix S4. Mean (weighted) observed life satisfaction levels over time.**

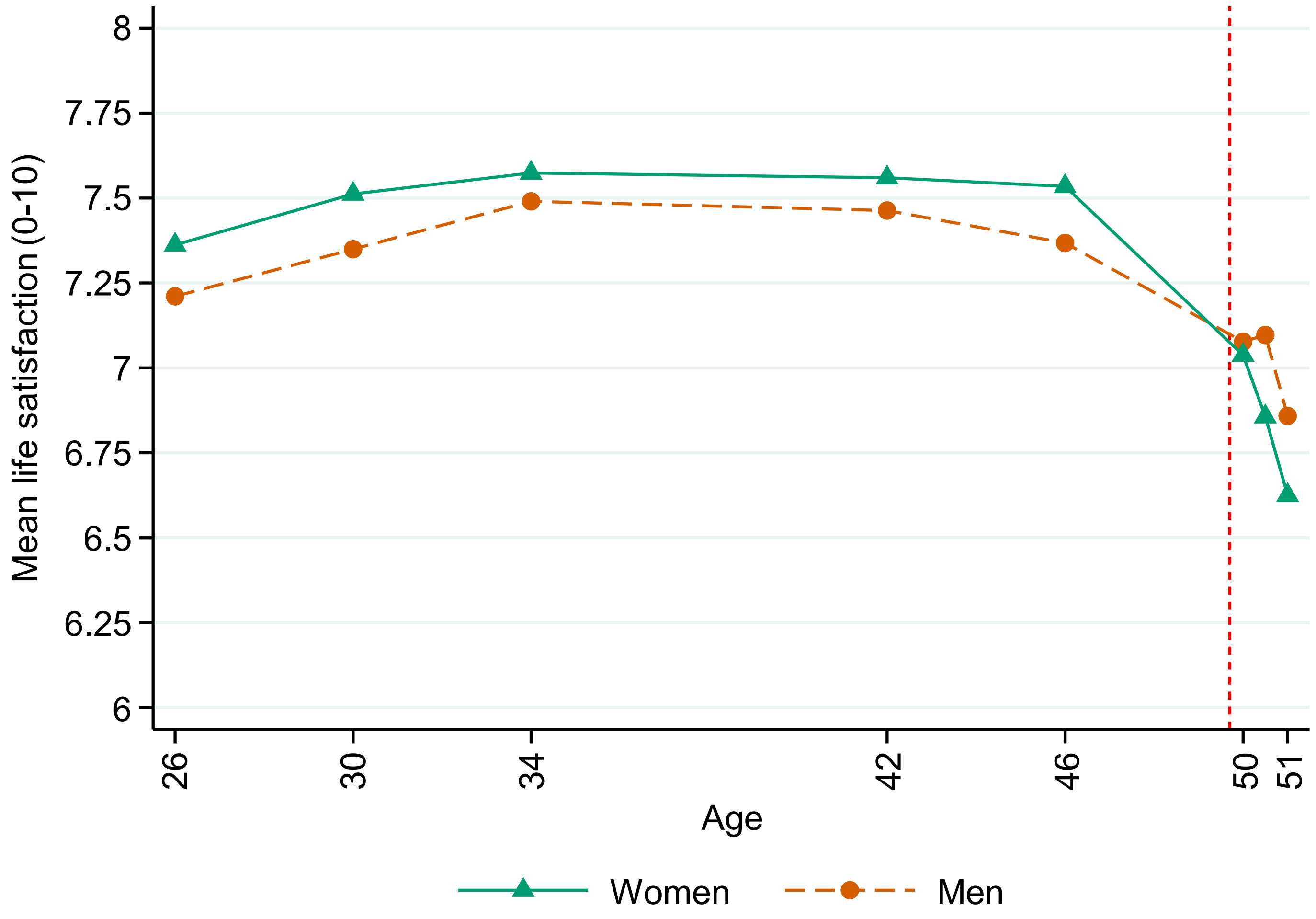

*Note.* The vertical red dashed line represents the pandemic onset.

**Appendix S5. Detailed information on the time-specific covariates (time use, financial situation, working from home, keyworker status, and dependent children or young people in the household) during the COVID-19 pandemic.**

| | Age 50 (May 2020) | | | | | | Age 50.5 (Sep/Oct 2020) | | | | | | Age 51 (Feb/Mar 2021) | | | | | |
|---|---|---|---|---|---|---|---|---|---|---|---|---|---|---|---|---|---|---|
| | Men (N=1599) | | Women (N=2309) | | Overall (N=3908) | | Men (N=2102) | | Women (N=2895) | | Overall (N=4997) | | Men (N=2312) | | Women (N=3132) | | Overall (N=5444) | |
| | N obs. | % | N obs. | % | N obs. | % | N obs. | % | N obs. | % | N obs. | % | N obs. | % | N obs. | % | N obs. | % |
| **Time spent working** | | | | | | | | | | | | | | | | | | |
| 0 hours | 423 | 26.5 | 798 | 34.6 | 1221 | 31.2 | 262 | 12.5 | 530 | 18.3 | 792 | 15.8 | 377 | 16.3 | 843 | 26.9 | 1220 | 22.4 |
| 1-8 hours | 786 | 49.2 | 1106 | 47.9 | 1892 | 48.4 | 1010 | 48.0 | 1515 | 52.3 | 2525 | 50.5 | 1157 | 50.0 | 1914 | 61.1 | 3071 | 56.4 |
| More than 8 hours | 378 | 23.6 | 383 | 16.6 | 761 | 19.5 | 728 | 34.6 | 747 | 25.8 | 1475 | 29.5 | 752 | 32.5 | 351 | 11.2 | 1103 | 20.3 |
| *Missing* | *12* | *0.8* | *22* | *1.0* | *34* | *0.9* | *102* | *4.9* | *103* | *3.6* | *205* | *4.1* | *26* | *1.1* | *24* | *0.8* | *50* | *0.9* |
| **Time spent volunteering** | | | | | | | | | | | | | | | | | | |
| 0 hours | 1492 | 93.3 | 2040 | 88.3 | 3532 | 90.4 | 1804 | 85.8 | 2486 | 85.9 | 4290 | 85.9 | | | | | | |
| 1+ hours | 95 | 5.9 | 247 | 10.7 | 342 | 8.8 | 196 | 9.3 | 306 | 10.6 | 502 | 10.0 | | | | | | |
| *Missing* | *12* | *0.8* | *22* | *1.0* | *34* | *0.9* | *102* | *4.9* | *103* | *3.6* | *205* | *4.1* | | | | | | |
| **Time spent home-schooling** | | | | | | | | | | | | | | | | | | |
| 0 hours | 1229 | 76.9 | 1676 | 72.6 | 2905 | 74.3 | 1744 | 83.0 | 2517 | 86.9 | 4261 | 85.3 | | | | | | |
| 1+ hours | 358 | 22.4 | 611 | 26.5 | 969 | 24.8 | 256 | 12.2 | 275 | 9.5 | 531 | 10.6 | | | | | | |
| *Missing* | *12* | *0.8* | *22* | *1.0* | *34* | *0.9* | *102* | *4.9* | *103* | *3.6* | *205* | *4.1* | | | | | | |
| **Time spent caring for children** | | | | | | | | | | | | | | | | | | |
| 0 hours | 1064 | 66.5 | 1580 | 68.4 | 2644 | 67.7 | 1273 | 60.6 | 1921 | 66.4 | 3194 | 63.9 | | | | | | |
| 1+ hours | 523 | 32.7 | 707 | 30.6 | 1230 | 31.5 | 727 | 34.6 | 871 | 30.1 | 1598 | 32.0 | | | | | | |
| *Missing* | *12* | *0.8* | *22* | *1.0* | *34* | *0.9* | *102* | *4.9* | *103* | *3.6* | *205* | *4.1* | | | | | | |
| **Time spent caring for others** | | | | | | | | | | | | | | | | | | |
| 0 hours | 1418 | 88.7 | 1907 | 82.6 | 3325 | 85.1 | 1805 | 85.9 | 2283 | 78.9 | 4088 | 81.8 | | | | | | |
| 1+ hours | 169 | 10.6 | 380 | 16.5 | 549 | 14.0 | 195 | 9.3 | 509 | 17.6 | 704 | 14.1 | | | | | | |
| *Missing* | *12* | *0.8* | *22* | *1.0* | *34* | *0.9* | *102* | *4.9* | *103* | *3.6* | *205* | *4.1* | | | | | | |
| **Time spent doing housework** | | | | | | | | | | | | | | | | | | |
| 0 hours | 294 | 18.4 | 102 | 4.4 | 396 | 10.1 | 196 | 9.3 | 70 | 2.4 | 266 | 5.3 | | | | | | |
| Up to 1 hour | 739 | 46.2 | 842 | 36.5 | 1581 | 40.5 | 1135 | 54.0 | 1040 | 35.9 | 2175 | 43.5 | | | | | | |
| More than 1 hour | 554 | 34.6 | 1343 | 58.2 | 1897 | 48.5 | 669 | 31.8 | 1682 | 58.1 | 2351 | 47.0 | | | | | | |
| *Missing* | *12* | *0.8* | *22* | *1.0* | *34* | *0.9* | *102* | *4.9* | *103* | *3.6* | *205* | *4.1* | | | | | | |
| **Financial situation compared to before the outbreak** | | | | | | | | | | | | | | | | | | |

| | | | | | | | | | | | | | | | | | | |
|---|---|---|---|---|---|---|---|---|---|---|---|---|---|---|---|---|---|---|
| Much worse off | 173 | 10.8 | 221 | 9.6 | 394 | 10.1 | 165 | 7.8 | 215 | 7.4 | 380 | 7.6 | 238 | 10.3 | 327 | 10.4 | 565 | 10.4 |
| Little worse off | 338 | 21.1 | 558 | 24.2 | 896 | 22.9 | 367 | 17.5 | 531 | 18.3 | 898 | 18.0 | 374 | 16.2 | 540 | 17.2 | 914 | 16.8 |
| About the same | 709 | 44.3 | 1027 | 44.5 | 1736 | 44.4 | 1053 | 50.1 | 1549 | 53.5 | 2602 | 52.1 | 961 | 41.6 | 1427 | 45.6 | 2388 | 43.9 |
| Little better off | 325 | 20.3 | 424 | 18.4 | 749 | 19.2 | 430 | 20.5 | 494 | 17.1 | 924 | 18.5 | 567 | 24.5 | 656 | 20.9 | 1223 | 22.5 |
| Much better off | 54 | 3.4 | 73 | 3.2 | 127 | 3.2 | 86 | 4.1 | 102 | 3.5 | 188 | 3.8 | 166 | 7.2 | 177 | 5.7 | 343 | 6.3 |
| *Missing* | *0* | *0.0* | *6* | *0.3* | *6* | *0.2* | *1* | *0.0* | *4* | *0.1* | *5* | *0.1* | *6* | *0.3* | *5* | *0.2* | *11* | *0.2* |
| **Works from home** | | | | | | | | | | | | | | | | | | |
| Never | 935 | 58.5 | 1406 | 60.9 | 2341 | 59.9 | 1209 | 57.5 | 1863 | 64.4 | 3072 | 61.5 | 1279 | 55.3 | 1888 | 60.3 | 3167 | 58.2 |
| Part or all of the time | 663 | 41.5 | 885 | 38.3 | 1548 | 39.6 | 890 | 42.3 | 1029 | 35.5 | 1919 | 38.4 | 1028 | 44.5 | 1243 | 39.7 | 2271 | 41.7 |
| *Missing* | *1* | *0.1* | *18* | *0.8* | *19* | *0.5* | *3* | *0.1* | *3* | *0.1* | *6* | *0.1* | *5* | *0.2* | *1* | *0.0* | *6* | *0.1* |
| **Keyworker** | | | | | | | | | | | | | | | | | | |
| No | 1071 | 67.0 | 1448 | 62.7 | 2519 | 64.5 | 1352 | 64.3 | 1678 | 58.0 | 3030 | 60.6 | 1424 | 61.6 | 1684 | 53.8 | 3108 | 57.1 |
| Yes | 525 | 32.8 | 843 | 36.5 | 1368 | 35.0 | 746 | 35.5 | 1212 | 41.9 | 1958 | 39.2 | 881 | 38.1 | 1444 | 46.1 | 2325 | 42.7 |
| *Missing* | *3* | *0.2* | *18* | *0.8* | *21* | *0.5* | *4* | *0.2* | *5* | *0.2* | *9* | *0.2* | *7* | *0.3* | *4* | *0.1* | *11* | *0.2* |
| **Dependent children or young people in the household** | | | | | | | | | | | | | | | | | | |
| No | 913 | 57.1 | 1381 | 59.8 | 2294 | 58.7 | 1271 | 60.5 | 1855 | 64.1 | 3126 | 62.6 | 1537 | 66.5 | 2158 | 68.9 | 3695 | 67.9 |
| Yes | 685 | 42.8 | 926 | 40.1 | 1611 | 41.2 | 826 | 39.3 | 1027 | 35.5 | 1853 | 37.1 | 761 | 32.9 | 959 | 30.6 | 1720 | 31.6 |
| *Missing* | *1* | *0.1* | *2* | *0.1* | *3* | *0.1* | *5* | *0.2* | *13* | *0.4* | *18* | *0.4* | *14* | *0.6* | *15* | *0.5* | *29* | *0.5* |

*Note.* Unweighted results. N obs: total number of observations.

**Appendix S6. Fit indices for the latent growth curve models estimated to identify the optimal functional form in the overall, women, and men samples.**

**Women**

| Model | Comp. model for $\Delta\chi^2$ | $\chi^2$ | df | Scaling correction factor | AIC | BIC | RMSEA | RMSEA LB | RMSEA UB | CFI | TLI | SRMR | $\Delta\chi^2$ | df | $\Delta\chi^2$ *p*-value |
|---|---|---|---|---|---|---|---|---|---|---|---|---|---|---|---|
| 1 No change / Intercepts-only | | 669.1 | 34 | 2.13 | 80093 | 80156 | 0.070 | 0.066 | 0.075 | 0.666 | 0.725 | 0.146 | | | |
| 2 Linear change | a | 342.1 | 31 | 2.09 | 79390 | 79471 | 0.051 | 0.047 | 0.056 | 0.837 | 0.852 | 0.071 | 279.24 | 3 | <0.001 |
| 3 Quadratic change | b | 122.5 | 27 | 2.12 | 78943 | 79049 | 0.031 | 0.025 | 0.036 | 0.95 | 0.948 | 0.050 | 241.21 | 4 | <0.001 |
| 4 Cubic change | c | 88.9 | 22 | 2.06 | 78876 | 79013 | 0.028 | 0.022 | 0.035 | 0.965 | 0.955 | 0.044 | 32.12 | 5 | <0.001 |
| 5 Piecewise: linear + linear | a | 72.3 | 27 | 2.10 | 78835 | 78941 | 0.021 | 0.015 | 0.027 | 0.976 | 0.975 | 0.038 | 567.01 | 7 | <0.001 |
| 6 Piecewise: quadratic + linear | e | 61.2 | 22 | 1.98 | 78814 | 78952 | 0.022 | 0.015 | 0.028 | 0.979 | 0.974 | 0.037 | 11.66 | 5 | 0.040 |
| **7 Piecewise: quadratic + quadratic** | **f** | **42.6** | **16** | **1.83** | **78783** | **78958** | **0.021** | **0.013** | **0.029** | **0.986** | **0.976** | **0.032** | **18.16** | **6** | **0.005** |
| 8 Free / latent basis | a | 145.5 | 25 | 2.08 | 78990 | 79108 | 0.036 | 0.030 | 0.041 | 0.937 | 0.929 | 0.048 | 494.75 | 9 | <0.001 |

**Men**

| Model | Comp. model for $\Delta\chi^2$ | $\chi^2$ | df | Scaling correction factor | AIC | BIC | RMSEA | RMSEA LB | RMSEA UB | CFI | TLI | SRMR | $\Delta\chi^2$ | df | $\Delta\chi^2$ *p*-value |
|---|---|---|---|---|---|---|---|---|---|---|---|---|---|---|---|
| 1 No change / Intercepts-only | | 350.7 | 34 | 2.20 | 55709 | 55769 | 0.056 | 0.051 | 0.061 | 0.813 | 0.846 | 0.124 | | | |
| 2 Linear change | a | 174.4 | 31 | 2.11 | 55310 | 55388 | 0.039 | 0.034 | 0.045 | 0.916 | 0.924 | 0.063 | 128.93 | 3 | <0.001 |
| 3 Quadratic change | b | 56.1 | 27 | 2.01 | 55064 | 55166 | 0.019 | 0.012 | 0.026 | 0.983 | 0.982 | 0.032 | 91.64 | 4 | <0.001 |
| 4 Cubic change | c | 27.9 | 22 | 1.86 | 55013 | 55145 | 0.010 | <0.001 | 0.019 | 0.997 | 0.996 | 0.019 | 22.80 | 5 | <0.001 |
| 5 Piecewise: linear + linear | a | 59.9 | 27 | 2.00 | 55071 | 55173 | 0.020 | 0.013 | 0.027 | 0.981 | 0.980 | 0.034 | 219.34 | 7 | <0.001 |
| 6 Piecewise: quadratic + linear | e | 34.1 | 22 | 1.81 | 55023 | 55155 | 0.014 | 0.001 | 0.022 | 0.993 | 0.991 | 0.023 | 20.48 | 5 | 0.001 |

| | | | | | | | | | | | | | | | |
|---|---|---|---|---|---|---|---|---|---|---|---|---|---|---|---|
| **7 Piecewise: quadratic + quadratic** | **f** | **18.8** | **16** | **1.94** | **55009** | **55177** | **0.008** | **<0.001** | **0.019** | **0.998** | **0.997** | **0.021** | **17.25** | **6** | **0.008** |
| 8 Free / latent basis | a | 105.1 | 25 | 2.00 | 55165 | 55279 | 0.033 | 0.027 | 0.039 | 0.953 | 0.947 | 0.039 | 203.71 | 9 | <0.001 |

**Overall**

| Model | Comp. model for $\Delta\chi^2$ | $\chi^2$ | df | Scaling correction factor | AIC | BIC | RMSEA | RMSEA LB | RMSEA UB | CFI | TLI | SRMR | $\Delta\chi^2$ | df | $\Delta\chi^2$ *p*-value |
|---|---|---|---|---|---|---|---|---|---|---|---|---|---|---|---|
| a) No change / Intercepts-only | | 837.2 | 34 | 2.33 | 134545 | 134613 | 0.059 | 0.056 | 0.063 | 0.758 | 0.801 | 0.130 | | | |
| b) Linear change | a | 416.3 | 31 | 2.23 | 133528 | 133616 | 0.043 | 0.039 | 0.047 | 0.884 | 0.895 | 0.059 | 303.96 | 3 | <0.001 |
| c) Quadratic change | b | 118.5 | 27 | 2.23 | 132870 | 132986 | 0.022 | 0.018 | 0.027 | 0.972 | 0.971 | 0.033 | 297.80 | 4 | <0.001 |
| d) Cubic change | c | 64.6 | 22 | 2.16 | 132756 | 132906 | 0.017 | 0.012 | 0.022 | 0.987 | 0.984 | 0.025 | 49.14 | 5 | <0.001 |
| e) Piecewise: linear + linear | a | 81.8 | 27 | 2.22 | 132788 | 132904 | 0.017 | 0.013 | 0.022 | 0.984 | 0.983 | 0.026 | 642.30 | 7 | <0.001 |
| f) Piecewise: quadratic + linear | e | 47.4 | 22 | 2.10 | 132716 | 132866 | 0.013 | 0.008 | 0.018 | 0.992 | 0.99 | 0.021 | 29.86 | 5 | <0.001 |
| **g) Piecewise: quadratic + quadratic** | **f** | **19.5** | **16** | **2.07** | **132669** | **132860** | **0.006** | **<0.001** | **0.013** | **0.999** | **0.998** | **0.014** | **27.14** | **6** | **<0.001** |
| h) Free / latent basis | a | 201.2 | 25 | 2.23 | 133060 | 133189 | 0.032 | 0.028 | 0.036 | 0.947 | 0.941 | 0.039 | 575.97 | 9 | <0.001 |

*Note*. Models estimated using non-response weights and full information maximum likelihood under MLR estimation. Favoured models are highlighted in boldface. AIC: Akaike Information Criterion; BIC: Bayesian Information Criterion; CFI: Comparative Fit Index; Comp. model for $\Delta\chi^2$: comparison model used in the Satorra-Bentler scaled robust chi-square difference test; df: degrees of freedom; $\Delta\chi^2$: chi-square difference test statistic; RMSEA: root mean square error of approximation (LB: lower bound; UB: upper bound); SRMR: standardised root mean square residual; TLI: Tucker-Lewis Index; $\chi^2$: chi-square statistic.

**Appendix S7. Fit indices for the multiple groups latent growth curve models estimated to identify the most parsimonious growth model across groups.**

| Model | Constraints | Comp. model for $\Delta\chi^2$ | $\chi^2$ | df | Scaling correction factor | AIC | BIC | RMSEA | RMSEA LB | RMSEA UP | CFI | TLI | SRMR | $\Delta\chi^2$ | df | $\Delta\chi^2$ *p*-value |
|---|---|---|---|---|---|---|---|---|---|---|---|---|---|---|---|---|
| a | Baseline / no constraints | | 60.7 | 32 | 1.88 | 133793 | 134175 | 0.016 | 0.010 | 0.022 | 0.992 | 0.986 | 0.028 | | | |
| b | Equal quad1 mean across groups | a | 61.1 | 33 | 1.90 | 133793 | 134168 | 0.016 | 0.009 | 0.022 | 0.992 | 0.987 | 0.028 | 0.78 | 1 | 0.377 |
| c | b + equal lin1 mean across groups | b | 61.4 | 34 | 1.92 | 133793 | 134161 | 0.015 | 0.009 | 0.022 | 0.992 | 0.987 | 0.028 | 0.70 | 1 | 0.403 |
| d | c + equal lin2 mean across groups | c | 61.8 | 35 | 1.91 | 133791 | 134152 | 0.015 | 0.009 | 0.021 | 0.993 | 0.988 | 0.028 | 0.10 | 1 | 0.752 |
| e | d + equal quad2 mean across groups | d | - | | | | | | | | | | | | | |
| f | d + quad2 variance fixed to zero in both groups | d | 68.1 | 45 | 1.91 | 133782 | 134076 | 0.012 | 0.006 | 0.018 | 0.994 | 0.992 | 0.029 | 6.30 | 10 | 0.790 |
| g | f + quad1 variance fixed to zero in both groups | f | 81.3 | 53 | 2.04 | 133803 | 134041 | 0.013 | 0.007 | 0.018 | 0.992 | 0.992 | 0.031 | 12.91 | 8 | 0.115 |
| h | g + lin1 variance fixed to zero in both groups | g | 310.3 | 59 | 2.23 | 134316 | 134514 | 0.035 | 0.032 | 0.039 | 0.93 | 0.934 | 0.079 | 134.61 | 6 | <0.001 |
| i | f + equal lin1 variance across groups | g | 80.1 | 54 | 2.07 | 133801 | 134032 | 0.012 | 0.006 | 0.017 | 0.993 | 0.993 | 0.031 | -0.01 | 1 | 1 |
| j | i + lin2 variance fixed to zero in both groups | i | 375.7 | 60 | 2.02 | 134383 | 134574 | 0.039 | 0.036 | 0.043 | 0.913 | 0.918 | 0.069 | 377.78 | 6 | <0.001 |
| k | i + equal lin2 variance across groups | i | 78.8 | 55 | 2.11 | 133799 | 134024 | 0.011 | 0.005 | 0.017 | 0.993 | 0.993 | 0.031 | 0.11 | 1 | 0.740 |
| **l** | **k + equal int variance across groups** | **k** | **80.9** | **56** | **2.13** | **133803** | **134021** | **0.011** | **0.005** | **0.017** | **0.993** | **0.993** | **0.036** | **1.87** | **1** | **0.172** |

*Note*. Models estimated using non-response weights and full information maximum likelihood under MLR estimation. Favoured model is highlighted in boldface. AIC: Akaike Information Criterion; BIC: Bayesian Information Criterion; CFI: Comparative Fit Index; Comp. model for $\Delta\chi^2$: comparison model used in the Satorra-Bentler scaled robust chi-square difference test; df: degrees of freedom; $\Delta\chi^2$: chi-square difference test statistic; int: intercept term; lin1: linear growth term for the first segment of the piecewise model; lin2: linear growth term for the second segment of the piecewise model; quad1: quadratic growth term for the first segment of the piecewise model; quad2: quadratic growth term for the second segment of the piecewise model; RMSEA: root mean square error of approximation (LB: lower bound; UB: upper bound); SRMR: standardised root mean square residual; TLI: Tucker-Lewis Index; $\chi^2$: chi-square statistic.

**Appendix S8. Results from the adjusted multiple group latent growth curve models (n=6,766).**

| | Adjusted for time-use variables | | | | | | Fully adjusted * | | | | | |
|---|---|---|---|---|---|---|---|---|---|---|---|---|
| | Women | | | Men | | | Women | | | Men | | |
| | Point estimate | 95% CI, LB | 95% CI, UB | Point estimate | 95% CI, LB | 95% CI, UB | Point estimate | 95% CI, LB | 95% CI, UB | Point estimate | 95% CI, LB | 95% CI, UB |
| **Means** | | | | | | | | | | | | |
| Intercept | 7.188 | 7.093 | 7.283 | 6.970 | 6.852 | 7.087 | 7.185 | 7.090 | 7.280 | 6.966 | 6.848 | 7.084 |
| Linear change, first segment | *0.042* | *0.026* | *0.057* | *0.042* | *0.026* | *0.057* | *0.042* | *0.027* | *0.058* | *0.042* | *0.027* | *0.058* |
| Quadratic change, first segment | *-0.002* | *-0.002* | *-0.001* | *-0.002* | *-0.002* | *-0.001* | *-0.002* | *-0.002* | *-0.001* | *-0.002* | *-0.002* | *-0.001* |
| Linear change, second segment | *0.266* | *-0.091* | *0.622* | *0.266* | *-0.091* | *0.622* | *0.265* | *-0.107* | *0.637* | *0.265* | *-0.107* | *0.637* |
| Quadratic change, second segment | -0.089 | -0.162 | -0.017 | -0.073 | -0.152 | 0.006 | -0.090 | -0.166 | -0.014 | -0.068 | -0.151 | 0.014 |
| **Variances** | | | | | | | | | | | | |
| Intercept | *1.716* | *1.479* | *1.954* | *1.716* | *1.479* | *1.954* | *1.720* | *1.481* | *1.960* | *1.720* | *1.481* | *1.960* |
| Linear change, first segment | *0.115* | *0.086* | *0.145* | *0.115* | *0.086* | *0.145* | *0.115* | *0.085* | *0.144* | *0.115* | *0.085* | *0.144* |
| Quadratic change, first segment | *0* | | | *0* | | | *0* | | | *0* | | |
| Linear change, second segment | *1.874* | *1.504* | *2.244* | *1.874* | *1.504* | *2.244* | *1.821* | *1.459* | *2.184* | *1.821* | *1.459* | *2.184* |
| Quadratic change, second segment | *0* | | | *0* | | | *0* | | | *0* | | |
| **Standardised covariances (correlations)** | | | | | | | | | | | | |
| Intercept x linear change, first segment | -0.351 | -0.476 | -0.226 | -0.318 | -0.449 | -0.188 | -0.350 | -0.476 | -0.224 | -0.316 | -0.446 | -0.185 |
| Intercept x linear change, second segment | -0.098 | -0.220 | 0.024 | -0.129 | -0.291 | 0.033 | -0.119 | -0.242 | 0.004 | -0.147 | -0.313 | 0.018 |
| Linear change first x second segment | -0.144 | -0.297 | 0.009 | -0.133 | -0.325 | 0.059 | -0.148 | -0.301 | 0.006 | -0.124 | -0.317 | 0.070 |
| **Standardised residual variances** | | | | | | | | | | | | |
| Age 26 | 0.554 | 0.498 | 0.610 | 0.547 | 0.487 | 0.607 | 0.554 | 0.498 | 0.610 | 0.547 | 0.488 | 0.606 |
| Age 30 | 0.561 | 0.515 | 0.608 | 0.512 | 0.453 | 0.572 | 0.561 | 0.515 | 0.608 | 0.511 | 0.451 | 0.571 |
| Age 34 | 0.545 | 0.500 | 0.591 | 0.477 | 0.417 | 0.537 | 0.543 | 0.497 | 0.589 | 0.476 | 0.416 | 0.536 |
| Age 42 | 0.543 | 0.482 | 0.603 | 0.502 | 0.448 | 0.557 | 0.543 | 0.482 | 0.603 | 0.500 | 0.445 | 0.554 |
| Age 46 | 0.398 | 0.325 | 0.471 | 0.378 | 0.300 | 0.457 | 0.400 | 0.327 | 0.472 | 0.380 | 0.303 | 0.457 |
| Age 50 | 0.418 | 0.348 | 0.487 | 0.268 | 0.220 | 0.315 | 0.416 | 0.359 | 0.474 | 0.269 | 0.221 | 0.317 |
| Age 50.5 | 0.281 | 0.240 | 0.322 | 0.292 | 0.234 | 0.350 | 0.285 | 0.245 | 0.324 | 0.288 | 0.231 | 0.346 |
| Age 51 | 0.339 | 0.289 | 0.389 | 0.260 | 0.210 | 0.311 | 0.326 | 0.280 | 0.373 | 0.254 | 0.205 | 0.303 |
| **Effects on life satisfaction at age 50** | | | | | | | | | | | | |
| Working 1-8 hours | 0.200 | -0.041 | 0.442 | 0.154 | -0.059 | 0.368 | 0.166 | -0.130 | 0.462 | 0.123 | -0.124 | 0.371 |
| Working more than 8 hours | 0.217 | -0.086 | 0.521 | -0.066 | -0.300 | 0.168 | 0.193 | -0.163 | 0.549 | -0.085 | -0.349 | 0.179 |
| Volunteering 1 or more hours | -0.174 | -0.432 | 0.084 | 0.091 | -0.246 | 0.429 | -0.160 | -0.410 | 0.091 | 0.081 | -0.249 | 0.411 |
| Home-schooling 1 or more hours | 0.148 | -0.102 | 0.398 | -0.036 | -0.249 | 0.178 | 0.247 | -0.050 | 0.543 | -0.069 | -0.298 | 0.161 |
| Caring for children 1 or more hours | -0.158 | -0.418 | 0.102 | -0.125 | -0.310 | 0.060 | -0.069 | -0.335 | 0.197 | -0.121 | -0.329 | 0.087 |

| | | | | | | | | | | | | |
|---|---|---|---|---|---|---|---|---|---|---|---|---|
| Caring for others 1 or more hours | 0.246 | -0.035 | 0.527 | 0.211 | -0.043 | 0.465 | 0.217 | -0.055 | 0.490 | 0.219 | -0.037 | 0.474 |
| Doing housework 1 hour | -0.127 | -0.473 | 0.219 | 0.118 | -0.088 | 0.323 | -0.166 | -0.532 | 0.199 | 0.117 | -0.090 | 0.324 |
| Doing housework 2 or more hours | -0.172 | -0.545 | 0.201 | -0.002 | -0.223 | 0.218 | -0.218 | -0.624 | 0.189 | -0.008 | -0.228 | 0.212 |
| Financial situation | | | | | | | 0.106 | -0.077 | 0.288 | 0.139 | 0.043 | 0.235 |
| Working from home | | | | | | | 0.181 | -0.016 | 0.377 | 0.059 | -0.153 | 0.272 |
| Keyworker status | | | | | | | 0.125 | -0.102 | 0.352 | -0.018 | -0.221 | 0.184 |
| Dependent CYP in the household | | | | | | | -0.080 | -0.374 | 0.214 | -0.065 | -0.289 | 0.158 |
| **Effects on life satisfaction at age 50.5** | | | | | | | | | | | | |
| Working 1-8 hours | 0.489 | 0.257 | 0.721 | -0.126 | -0.420 | 0.168 | 0.338 | 0.105 | 0.571 | -0.197 | -0.497 | 0.103 |
| Working more than 8 hours | 0.389 | 0.135 | 0.643 | -0.289 | -0.623 | 0.046 | 0.251 | -0.005 | 0.508 | -0.309 | -0.640 | 0.022 |
| Volunteering 1 or more hours | 0.101 | -0.106 | 0.308 | 0.268 | 0.010 | 0.527 | 0.096 | -0.110 | 0.302 | 0.228 | -0.030 | 0.487 |
| Home-schooling 1 or more hours | -0.153 | -0.420 | 0.113 | -0.087 | -0.422 | 0.249 | -0.190 | -0.464 | 0.084 | -0.144 | -0.500 | 0.212 |
| Caring for children 1 or more hours | 0.133 | -0.023 | 0.289 | 0.131 | -0.069 | 0.331 | 0.120 | -0.052 | 0.293 | 0.100 | -0.160 | 0.360 |
| Caring for others 1 or more hours | -0.190 | -0.389 | 0.008 | 0.236 | -0.097 | 0.570 | -0.200 | -0.403 | 0.002 | 0.285 | -0.059 | 0.629 |
| Doing housework 1 hour | -0.176 | -0.456 | 0.104 | 0.349 | 0.078 | 0.620 | -0.203 | -0.483 | 0.077 | 0.352 | 0.075 | 0.629 |
| Doing housework 2 or more hours | -0.150 | -0.422 | 0.121 | 0.230 | -0.104 | 0.564 | -0.160 | -0.432 | 0.112 | 0.249 | -0.088 | 0.586 |
| Financial situation | | | | | | | 0.115 | 0.019 | 0.212 | 0.148 | 0.041 | 0.255 |
| Working from home | | | | | | | 0.376 | 0.201 | 0.551 | 0.231 | 0.014 | 0.448 |
| Keyworker status | | | | | | | 0.171 | -0.033 | 0.374 | -0.255 | -0.493 | -0.017 |
| Dependent CYP in the household | | | | | | | 0.007 | -0.255 | 0.268 | -0.038 | -0.315 | 0.238 |
| **Effects on life satisfaction at age 51** | | | | | | | | | | | | |
| Working 1-8 hours | 0.245 | 0.005 | 0.486 | 0.282 | -0.053 | 0.618 | 0.286 | -0.022 | 0.595 | 0.182 | -0.190 | 0.554 |
| Working more than 8 hours | 0.125 | -0.189 | 0.439 | 0.296 | -0.043 | 0.635 | 0.185 | -0.191 | 0.561 | 0.204 | -0.171 | 0.578 |
| Financial situation | | | | | | | 0.225 | 0.123 | 0.326 | 0.181 | 0.083 | 0.278 |
| Working from home | | | | | | | 0.060 | -0.164 | 0.284 | -0.008 | -0.264 | 0.247 |
| Keyworker status | | | | | | | -0.050 | -0.300 | 0.201 | 0.068 | -0.168 | 0.304 |
| Dependent CYP in the household | | | | | | | 0.100 | -0.209 | 0.409 | -0.138 | -0.424 | 0.148 |

*Note.* Models estimated using non-response weights and full information maximum likelihood under MLR estimation. Estimates in italics are constrained to be equal across groups. CI: confidence interval; CYP: children or young people; LB: lower bound; UB: upper bound. Effects of time use variables are estimated with no time spent in that activity as a reference category. * Adjusted for financial situation, working-from-home and keyworker status, and presence in the household of dependent children or young people aged up to 16 years old.

**Appendix S9. Results from the Wald tests analysing the difference in the impact of time-use variables on life satisfaction across women and men, based on fully adjusted multiple group latent growth curve models (n=6,766).**

| | Point estimate | 95% CI, LB | 95% CI, UB | p-value |
|---|---|---|---|---|
| **Difference in effect on life satisfaction at age 50** | | | | |
| Working 1-8 hours | 0.043 | -0.336 | 0.421 | 0.824 |
| Working more than 8 hours | 0.278 | -0.157 | 0.714 | 0.211 |
| Volunteering 1 or more hours | -0.241 | -0.654 | 0.173 | 0.254 |
| Home-schooling 1 or more hours | 0.315 | -0.060 | 0.690 | 0.099 |
| Caring for children 1 or more hours | 0.052 | -0.286 | 0.390 | 0.763 |
| Caring for others 1 or more hours | -0.001 | -0.374 | 0.371 | 0.994 |
| Doing housework 1 hour | -0.283 | -0.674 | 0.107 | 0.155 |
| Doing housework 2 or more hours | -0.209 | -0.636 | 0.217 | 0.336 |
| **Difference in effect on life satisfaction at age 50.5** | | | | |
| Working 1-8 hours | 0.535 | 0.156 | 0.914 | 0.006 |
| Working more than 8 hours | 0.560 | 0.143 | 0.978 | 0.009 |
| Volunteering 1 or more hours | -0.132 | -0.463 | 0.198 | 0.433 |
| Home-schooling 1 or more hours | -0.046 | -0.495 | 0.403 | 0.841 |
| Caring for children 1 or more hours | 0.020 | -0.293 | 0.334 | 0.898 |
| Caring for others 1 or more hours | -0.486 | -0.884 | -0.087 | 0.017 |
| Doing housework 1 hour | -0.555 | -0.956 | -0.155 | 0.007 |
| Doing housework 2 or more hours | -0.409 | -0.855 | 0.037 | 0.072 |
| **Difference in effect on life satisfaction at age 51** | | | | |
| Working 1-8 hours | 0.104 | -0.366 | 0.574 | 0.664 |
| Working more than 8 hours | -0.019 | -0.535 | 0.497 | 0.943 |

*Note.* Results based on models estimated using non-response weights and full information maximum likelihood under MLR estimation and adjusted for financial situation, working-from-home and keyworker status, and presence in the household of dependent children or young people aged up to 16 years old. CI: confidence interval; LB: lower bound; UB: upper bound. Effects of time use variables are estimated with no time spent in that activity as a reference category. Wald tests estimate the statistical significance of the null hypothesis of $estimate_{women}$ - $estimate_{men}$ = 0. Therefore, a positive point estimate indicates that the variable has a more positive or less negative effect on women's life satisfaction, whereas a negative point estimate indicates that the variable has a more positive or less negative effect on men's life satisfaction.

**Appendix S10. Results from the multiple group latent growth curve models adjusted for interview mode (n=6,766).**

| | Adjusted for interview mode only | | | | | | Fully adjusted * | | | | | |
|---|---|---|---|---|---|---|---|---|---|---|---|---|
| | Women | | | Men | | | Women | | | Men | | |
| | **Point estimate** | **95% CI, LB** | **95% CI, UB** | **Point estimate** | **95% CI, LB** | **95% CI, UB** | **Point estimate** | **95% CI, LB** | **95% CI, UB** | **Point estimate** | **95% CI, LB** | **95% CI, UB** |
| **Means** | | | | | | | | | | | | |
| Intercept | 7.195 | 7.103 | 7.288 | 6.978 | 6.863 | 7.093 | 7.188 | 7.093 | 7.283 | 6.966 | 6.848 | 7.085 |
| Linear change, first segment | *0.041* | *0.026* | *0.057* | *0.041* | *0.026* | *0.057* | *0.041* | *0.026* | *0.057* | *0.041* | *0.026* | *0.057* |
| Quadratic change, first segment | *-0.002* | *-0.002* | *-0.001* | *-0.002* | *-0.002* | *-0.001* | *-0.002* | *-0.002* | *-0.001* | *-0.002* | *-0.002* | *-0.001* |
| Linear change, second segment | *0.248* | *0.156* | *0.341* | *0.248* | *0.156* | *0.341* | *0.313* | *-0.051* | *0.677* | *0.313* | *-0.051* | *0.677* |
| Quadratic change, second segment | -0.082 | -0.102 | -0.063 | -0.061 | -0.081 | -0.041 | -0.106 | -0.180 | -0.032 | -0.079 | -0.160 | 0.002 |
| **Variances** | | | | | | | | | | | | |
| Intercept | *1.698* | *1.465* | *1.932* | *1.698* | *1.465* | *1.932* | *1.720* | *1.481* | *1.960* | *1.720* | *1.481* | *1.960* |
| Linear change, first segment | *0.116* | *0.086* | *0.147* | *0.116* | *0.086* | *0.147* | *0.116* | *0.086* | *0.146* | *0.116* | *0.086* | *0.146* |
| Quadratic change, first segment | *0* | | | *0* | | | *0* | | | *0* | | |
| Linear change, second segment | *1.859* | *1.480* | *2.238* | *1.859* | *1.480* | *2.238* | *1.801* | *1.427* | *2.175* | *1.801* | *1.427* | *2.175* |
| Quadratic change, second segment | *0* | | | *0* | | | *0* | | | *0* | | |
| **Standardised covariances (correlations)** | | | | | | | | | | | | |
| Intercept x linear change, first segment | -0.346 | -0.473 | -0.219 | -0.326 | -0.456 | -0.197 | -0.347 | -0.475 | -0.219 | -0.323 | -0.453 | -0.193 |
| Intercept x linear change, second segment | -0.093 | -0.211 | 0.026 | -0.103 | -0.267 | 0.060 | -0.123 | -0.241 | -0.004 | -0.147 | -0.313 | 0.019 |
| Linear change first x second segment | -0.117 | -0.288 | 0.053 | -0.132 | -0.326 | 0.062 | -0.128 | -0.299 | 0.043 | -0.118 | -0.312 | 0.077 |
| **Standardised residual variances** | | | | | | | | | | | | |
| Age 26 | 0.556 | 0.499 | 0.613 | 0.550 | 0.489 | 0.611 | 0.554 | 0.498 | 0.610 | 0.547 | 0.488 | 0.605 |
| Age 30 | 0.564 | 0.518 | 0.610 | 0.517 | 0.457 | 0.577 | 0.561 | 0.515 | 0.607 | 0.512 | 0.452 | 0.572 |
| Age 34 | 0.547 | 0.501 | 0.593 | 0.483 | 0.422 | 0.545 | 0.542 | 0.496 | 0.588 | 0.479 | 0.419 | 0.539 |
| Age 42 | 0.544 | 0.483 | 0.604 | 0.507 | 0.452 | 0.562 | 0.542 | 0.482 | 0.603 | 0.500 | 0.446 | 0.555 |
| Age 46 | 0.390 | 0.316 | 0.464 | 0.382 | 0.300 | 0.464 | 0.391 | 0.318 | 0.464 | 0.381 | 0.305 | 0.457 |
| Age 50 | 0.411 | 0.337 | 0.484 | 0.272 | 0.222 | 0.323 | 0.409 | 0.348 | 0.470 | 0.270 | 0.222 | 0.318 |
| Age 50.5 | 0.298 | 0.256 | 0.340 | 0.285 | 0.223 | 0.348 | 0.281 | 0.238 | 0.324 | 0.289 | 0.231 | 0.347 |
| Age 51 | 0.332 | 0.282 | 0.382 | 0.273 | 0.221 | 0.325 | 0.322 | 0.277 | 0.367 | 0.254 | 0.206 | 0.301 |
| **Effects on life satisfaction at age 50** | | | | | | | | | | | | |
| Working 1-8 hours | | | | | | | 0.161 | -0.132 | 0.453 | 0.118 | -0.129 | 0.365 |
| Working more than 8 hours | | | | | | | 0.183 | -0.169 | 0.536 | -0.088 | -0.353 | 0.177 |
| Volunteering 1 or more hours | | | | | | | -0.169 | -0.419 | 0.080 | 0.090 | -0.242 | 0.421 |
| Home-schooling 1 or more hours | | | | | | | 0.241 | -0.060 | 0.541 | -0.070 | -0.299 | 0.159 |

| | | | | | | | | | | | | |
|---|---|---|---|---|---|---|---|---|---|---|---|---|
| Caring for children 1 or more hours | | | | | | | -0.069 | -0.335 | 0.197 | -0.146 | -0.365 | 0.074 |
| Caring for others 1 or more hours | | | | | | | 0.215 | -0.056 | 0.486 | 0.217 | -0.043 | 0.477 |
| Doing housework 1 hour | | | | | | | -0.148 | -0.501 | 0.205 | 0.106 | -0.103 | 0.314 |
| Doing housework 2 or more hours | | | | | | | -0.188 | -0.571 | 0.196 | -0.014 | -0.234 | 0.206 |
| Financial situation | | | | | | | 0.117 | -0.060 | 0.295 | 0.136 | 0.039 | 0.232 |
| Working from home | | | | | | | 0.180 | -0.015 | 0.375 | 0.049 | -0.160 | 0.259 |
| Keyworker status | | | | | | | 0.106 | -0.126 | 0.338 | -0.021 | -0.221 | 0.180 |
| Dependent CYP in the household | | | | | | | -0.068 | -0.363 | 0.228 | -0.046 | -0.269 | 0.177 |
| **Effects on life satisfaction at age 50.5** | | | | | | | | | | | | |
| Working 1-8 hours | | | | | | | 0.333 | 0.102 | 0.565 | -0.193 | -0.490 | 0.104 |
| Working more than 8 hours | | | | | | | 0.247 | -0.008 | 0.501 | -0.302 | -0.631 | 0.027 |
| Volunteering 1 or more hours | | | | | | | 0.085 | -0.120 | 0.291 | 0.216 | -0.038 | 0.471 |
| Home-schooling 1 or more hours | | | | | | | -0.157 | -0.432 | 0.118 | -0.157 | -0.518 | 0.204 |
| Caring for children 1 or more hours | | | | | | | 0.116 | -0.057 | 0.290 | 0.064 | -0.186 | 0.314 |
| Caring for others 1 or more hours | | | | | | | -0.204 | -0.402 | -0.005 | 0.283 | -0.059 | 0.624 |
| Doing housework 1 hour | | | | | | | -0.160 | -0.428 | 0.108 | 0.376 | 0.100 | 0.651 |
| Doing housework 2 or more hours | | | | | | | -0.112 | -0.371 | 0.147 | 0.271 | -0.064 | 0.606 |
| Financial situation | | | | | | | 0.109 | 0.012 | 0.205 | 0.157 | 0.053 | 0.261 |
| Working from home | | | | | | | 0.375 | 0.200 | 0.550 | 0.216 | 0.002 | 0.430 |
| Keyworker status | | | | | | | 0.181 | -0.027 | 0.389 | -0.255 | -0.490 | -0.019 |
| Dependent CYP in the household | | | | | | | 0.027 | -0.239 | 0.292 | -0.026 | -0.298 | 0.246 |
| **Effects on life satisfaction at age 51** | | | | | | | | | | | | |
| Working 1-8 hours | | | | | | | 0.329 | 0.026 | 0.632 | 0.205 | -0.164 | 0.573 |
| Working more than 8 hours | | | | | | | 0.231 | -0.142 | 0.603 | 0.234 | -0.137 | 0.605 |
| Financial situation | | | | | | | 0.213 | 0.114 | 0.313 | 0.163 | 0.064 | 0.262 |
| Working from home | | | | | | | 0.081 | -0.145 | 0.306 | -0.019 | -0.269 | 0.230 |
| Keyworker status | | | | | | | -0.057 | -0.314 | 0.200 | 0.081 | -0.151 | 0.312 |
| Dependent CYP in the household | | | | | | | 0.119 | -0.200 | 0.437 | -0.147 | -0.423 | 0.128 |
| Phone interview | 1.264 | -1.150 | 3.678 | 0.115 | -0.809 | 1.040 | 1.645 | -0.610 | 3.900 | -0.027 | -1.266 | 1.213 |

*Note.* Models estimated using non-response weights and full information maximum likelihood under MLR estimation. Estimates in italics are constrained to be equal across groups. CI: confidence interval; CYP: children or young people; LB: lower bound; UB: upper bound. Effects of time use variables are estimated with no time spent in that activity as a reference category. * Adjusted for financial situation, working-from-home and keyworker status, presence in the household of dependent children or young people aged up to 16 years old, and interview mode.

**Appendix S11. Results from the multiple group latent growth curve models including lagged effects of the time-specific variables (n=6,766).**

| | Women | | | Men | | |
|---|---|---|---|---|---|---|
| | **Point estimate** | **95% CI, LB** | **95% CI, UB** | **Point estimate** | **95% CI, LB** | **95% CI, UB** |
| **Means** | | | | | | |
| Intercept | 7.187 | 7.093 | 7.281 | 6.969 | 6.851 | 7.087 |
| Linear change, first segment | *0.041* | *0.026* | *0.057* | *0.041* | *0.026* | *0.057* |
| Quadratic change, first segment | *-0.002* | *-0.002* | *-0.001* | *-0.002* | *-0.002* | *-0.001* |
| Linear change, second segment | *0.133* | *-0.164* | *0.430* | *0.133* | *-0.164* | *0.430* |
| Quadratic change, second segment | -0.061 | -0.132 | 0.011 | -0.036 | -0.109 | 0.038 |
| **Variances** | | | | | | |
| Intercept | *1.712* | *1.475* | *1.949* | *1.712* | *1.475* | *1.949* |
| Linear change, first segment | *0.116* | *0.086* | *0.147* | *0.116* | *0.086* | *0.147* |
| Quadratic change, first segment | *0* | | | *0* | | |
| Linear change, second segment | *1.918* | *1.513* | *2.323* | *1.918* | *1.513* | *2.323* |
| Quadratic change, second segment | *0* | | | *0* | | |
| **Standardised covariances (correlations)** | | | | | | |
| Intercept x linear change, first segment | -0.349 | -0.477 | -0.221 | -0.324 | -0.453 | -0.194 |
| Intercept x linear change, second segment | -0.081 | -0.199 | 0.037 | -0.097 | -0.260 | 0.067 |
| Linear change first x second segment | -0.103 | -0.273 | 0.066 | -0.145 | -0.334 | 0.044 |
| **Standardised residual variances** | | | | | | |
| Age 26 | 0.554 | 0.498 | 0.610 | 0.550 | 0.489 | 0.610 |
| Age 30 | 0.563 | 0.516 | 0.609 | 0.514 | 0.455 | 0.574 |
| Age 34 | 0.545 | 0.499 | 0.590 | 0.479 | 0.419 | 0.539 |
| Age 42 | 0.543 | 0.482 | 0.604 | 0.503 | 0.449 | 0.557 |
| Age 46 | 0.393 | 0.319 | 0.467 | 0.379 | 0.301 | 0.457 |
| Age 50 | 0.400 | 0.336 | 0.464 | 0.269 | 0.220 | 0.318 |
| Age 50.5 | 0.278 | 0.237 | 0.318 | 0.281 | 0.222 | 0.339 |
| Age 51 | 0.339 | 0.292 | 0.386 | 0.268 | 0.218 | 0.318 |
| **Lagged effects on life satisfaction at age 50.5 (from age 50)** | | | | | | |
| Working 1-8 hours | -0.057 | -0.288 | 0.175 | -0.163 | -0.435 | 0.109 |
| Working more than 8 hours | -0.101 | -0.384 | 0.181 | -0.095 | -0.376 | 0.187 |
| Volunteering 1 or more hours | 0.129 | -0.091 | 0.349 | 0.375 | 0.004 | 0.747 |
| Home-schooling 1 or more hours | -0.096 | -0.338 | 0.146 | 0.054 | -0.216 | 0.325 |
| Caring for children 1 or more hours | 0.223 | -0.015 | 0.461 | 0.090 | -0.212 | 0.393 |

| | | | | | | |
|---|---|---|---|---|---|---|
| Caring for others 1 or more hours | -0.088 | -0.308 | 0.132 | 0.030 | -0.341 | 0.402 |
| Doing housework 1 hour | 0.146 | -0.131 | 0.423 | 0.068 | -0.183 | 0.318 |
| Doing housework 2 or more hours | 0.158 | -0.133 | 0.449 | 0.041 | -0.215 | 0.297 |
| Financial situation | 0.011 | -0.112 | 0.134 | -0.052 | -0.161 | 0.056 |
| Working from home | 0.034 | -0.146 | 0.214 | 0.128 | -0.079 | 0.336 |
| Keyworker status | 0.041 | -0.151 | 0.233 | -0.190 | -0.419 | 0.040 |
| Dependent CYP in the household | | | | | | |
| **Lagged effects on life satisfaction at age 51 (from age 50.5)** | | | | | | |
| Working 1-8 hours | -0.280 | -0.537 | -0.023 | 0.189 | -0.156 | 0.534 |
| Working more than 8 hours | -0.252 | -0.537 | 0.033 | 0.289 | -0.060 | 0.638 |
| Volunteering 1 or more hours | 0.105 | -0.139 | 0.349 | -0.077 | -0.364 | 0.209 |
| Home-schooling 1 or more hours | 0.101 | -0.211 | 0.412 | 0.114 | -0.234 | 0.462 |
| Caring for children 1 or more hours | -0.067 | -0.268 | 0.133 | -0.020 | -0.266 | 0.226 |
| Caring for others 1 or more hours | 0.106 | -0.141 | 0.353 | -0.212 | -0.536 | 0.113 |
| Doing housework 1 hour | 0.211 | -0.188 | 0.611 | -0.266 | -0.572 | 0.039 |
| Doing housework 2 or more hours | 0.214 | -0.192 | 0.620 | -0.266 | -0.630 | 0.097 |
| Financial situation | 0.013 | -0.117 | 0.142 | -0.073 | -0.179 | 0.033 |
| Working from home | -0.210 | -0.400 | -0.019 | -0.171 | -0.355 | 0.012 |
| Keyworker status | -0.025 | -0.231 | 0.182 | 0.052 | -0.136 | 0.241 |
| Dependent CYP in the household | 0.090 | -0.171 | 0.351 | 0.049 | -0.199 | 0.297 |
| **Concurrent effects on life satisfaction at age 51** | | | | | | |
| Phone interview | 1.588 | -0.540 | 3.716 | 0.165 | -1.140 | 1.471 |

*Note.* Models estimated using non-response weights and full information maximum likelihood under MLR estimation. Estimates in italics are constrained to be equal across groups. CI: confidence interval; CYP: children or young people; LB: lower bound; UB: upper bound. Effects of time use variables are estimated with no time spent in that activity as a reference category.
